# Evolutionary Measures Show that Recurrence of DCIS is Distinct from Progression to Breast Cancer

**DOI:** 10.1101/2024.08.15.24311949

**Authors:** Angelo Fortunato, Diego Mallo, Luis Cisneros, Lorraine M. King, Aziz Khan, Christina Curtis, Marc D. Ryser, Joseph Y. Lo, Allison Hall, Jeffrey R. Marks, E. Shelley Hwang, Carlo C. Maley

## Abstract

Progression from pre-cancers like ductal carcinoma *in situ* (DCIS) to invasive disease (cancer) is driven by somatic evolution and is altered by clinical interventions. We hypothesized that genetic and/or phenotypic intra-tumor heterogeneity would predict clinical outcomes for DCIS since it serves as the substrate for natural selection among cells. We profiled two samples from two geographically distinct foci from each DCIS in both cross-sectional (N = 119) and longitudinal cohorts (N = 224), with whole exome sequencing, low-pass whole genome sequencing, and a panel of immunohistochemical markers. In the longitudinal cohorts, the only statistically significant predictors of time to non-invasive DCIS recurrence were the combination of treatment (lumpectomy only vs mastectomy or lumpectomy with radiation, HR = 12.13, *p* = 0.003, Wald test with FDR correction), ER status (HR = 0.16 for ER+ compared to ER-, *p* = 0.0045), and divergence in SNVs between the two samples (HR = 1.33 per 10% divergence, *p* = 0.018). SNV divergence also distinguished between pure DCIS and DCIS synchronous with invasive disease in the cross-sectional cohort. In contrast, the only statistically significant predictors of time to progression to invasive disease were the combination of the width of the surgical margin (HR = 0.67 per mm, *p* = 0.043) and the number of mutations that were detectable at high allele frequencies (HR = 1.30 per 10 SNVs, *p* = 0.02). These results imply that recurrence with DCIS is a clinical and biological process different from invasive progression.

**Significance:** Evolutionary measures of breast pre-cancers associate with local recurrence after surgery, as well as progression to cancer. Recurrence and progression are different biological processes impacted differently by clinical interventions.

## Introduction

The improvement of radiological techniques and preventive screening of breast cancer conducted on a large scale makes it possible to identify mammary gland neoplasms at an early stage of development, when they are still confined within the glandular ducts. This neoplasm is termed ductal carcinoma *in situ* (DCIS) (1). Estimates from several natural history studies of DCIS indicate that 20-30% will progress to invasive cancer without definitive surgical treatment (2,3), implying that as many as 70% of patients who have surgery for DCIS may not derive benefit.

The ability to recognize which pre-cancerous tumors are likely to progress to invasive cancer is of great importance because it would identify high-risk patients for surgical, pharmacological, and/or radiation treatment. In contrast, low-risk patients could be managed by watchful waiting, avoiding the unnecessary harms and side effects associated with these therapies (4). Furthermore, selecting patients most at risk would facilitate reallocating healthcare resources to those who would benefit most from treatment.

Evolutionary mechanisms drive tumor progression (5). The impairment of control mechanisms of genetic integrity (6) accelerates the accumulation of new genetic alterations in cancer cells (7). The combination of these alterations in an increasing number of clones represents a critical factor in tumor progression, as these clones constitute the substrate upon which selection acts (8). The identification of mutations and the level of genetic (and phenotypic) heterogeneity have been shown to be associated with the risk of tumor progression in other pre-cancers, like Barrett’s esophagus (9–11). The higher the number of mutations and the greater the intratumor genetic heterogeneity, the higher the risk of developing clones that are cancerous, metastatic, and treatment-resistant (12–16).

It is challenging to integrate the combined effect of many mutations and genetic alterations that act simultaneously in cancer cells (17). Investigating the number of mutations and the level of heterogeneity allows us to introduce a quantitative parameter independent of the functional consequences of specific combinations of mutations, serving as a surrogate measure of the degree of evolvability of the neoplastic cells (18,19).

Both genetic and phenotypic heterogeneity can be measured by comparing different regions of the same tumor, ideally through analysis of longitudinal cohorts with linked clinical outcomes. Such studies often necessitate analysis of archival formalin-fixed paraffin-embedded (FFPE) samples, which is challenging due to partial degradation of the DNA, FFPE-induced artifacts, which manifest as sequence alterations, and low yield of nucleic acids from a limited number of sections. We recently published a workflow that overcomes these challenges, enabling the assessment of measures of genetic divergence between regions of the same tumor (20). This work aimed to test the hypothesis that genetic and phenotypic heterogeneity within DCIS can predict the recurrence of DCIS and/or progression to invasive ductal carcinoma (IDC).

## Materials and Methods

### Experimental design

We performed two observational studies (Fig. 1, Table 1) to study DCIS progression. In a cross-sectional study (Fig. 1A), we compared DCIS samples from patients with DCIS only (*Pure DCIS,* n = 58) versus DCIS samples from patients with synchronous invasive ductal carcinoma (*Synchronous DCIS*, n = 61). In a longitudinal case-control study (Fig. 1B), we collected samples from patients with primary DCIS who were treated and then followed until they were diagnosed with an IDC recurrence (*Progressors*, n = 56), were diagnosed with a DCIS-only recurrence (*Recurrents*, n = 69), or did not recur within their follow-up time (*Nonrecurrents,* n = 99, minimum five years). We calculated the median follow-up time using the reverse Kaplan-Meier method (21). In both cohorts, we characterized the genotype and phenotype of two DCIS regions per patient. All samples came from different FFPE blocks or were separated by at least 8mm. For some progressors, we also obtained a subsequent IDC sample. The Institutional Review Board (IRB) of Duke University Medical Center approved this study, and a waiver of consent was obtained according to the approved protocol.

**Figure 1.**
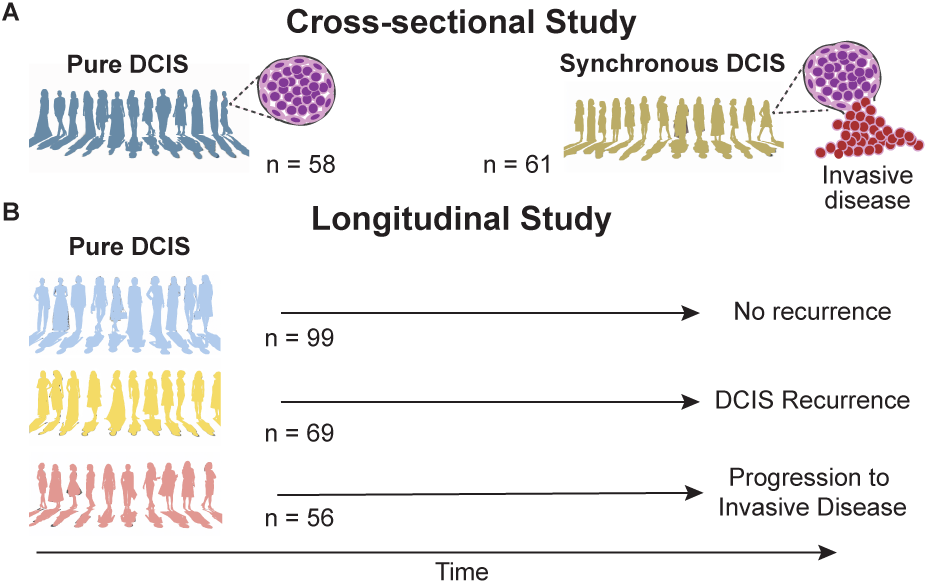
Schematic of the two study designs. **A**: Cross-sectional study: Synchronous DCIS tumors are presumed to have evolved from pure DCIS that existed before the progression of the synchronous IDC. In patients with synchronous DCIS, only the DCIS component was sampled and assayed unless otherwise specified. **B**: Longitudinal case-control study: pure-DCIS samples from patients treated and followed up for at least five years or until they progress or recur. n: number of patients per cohort.

**Table 1.**
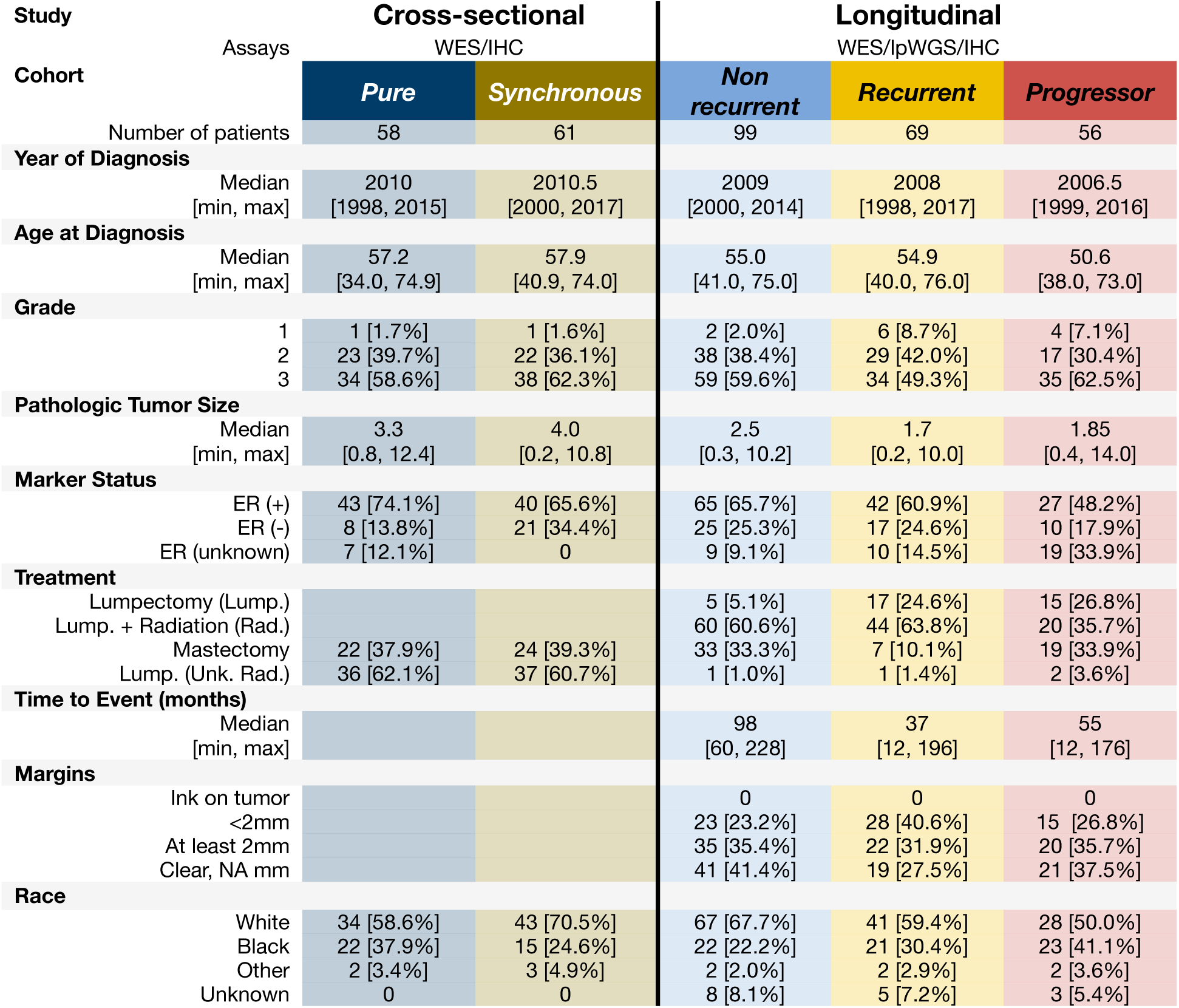
Patient Cohorts with WES data, lpWGS, or IHC data. Unk. = Unknown.

### Clinical specimens

We classified breast tumors according to the World Health Organization (WHO) criteria (22). We graded the IDC and DCIS samples according to the Nottingham grading system (23) or recommendations from the Consensus Conference on DCIS classification (24), respectively.

All samples were obtained from formalin-fixed paraffin-embedded (FFPE) breast tissue blocks. Cases from the cross-sectional studies were obtained from the Duke Pathology archives. Cases from the longitudinal study were obtained from Translational Breast Cancer Research Consortium (TBCRC) sites, a national multi-center consortium of cancer centers that treat breast cancer patients. All cases underwent detailed pathology review (AH) for histologic features and case eligibility.

### DNA extraction and sequencing

The DNA extraction, sequencing, and data processing protocol has been previously reported (20). For each neoplastic sample, we extracted the DNA from multiple serial archival FFPE tissue block sections after macro-dissecting the areas of interest. To estimate the germline sequence, we also extracted DNA from either distant benign breast tissue or a benign lymph node. The study pathologist confirmed the presence of ≥70% neoplastic cells in the microdissected areas of neoplastic samples and their absence from control samples.

After DNA extraction, hybrid capture was performed using two targeted panels (all exons of the 83 genes in the breast cancer gene panel and the human exome), and the multiplexed libraries were sequenced using either an Illumina HiSeq with 4-channel chemistry (cross-sectional study) or a NovaSeq 6000 machine with 2-channel chemistry (longitudinal study). After alignment to the Genome Reference Consortium Human Build 37 and marking duplicates, we obtained a mean de-duplicated depth of 115.9 ± 52.2 (SD). The resulting BAM files were the input data for our SNV calling and heterogeneity calculation pipeline. We discarded samples with less than 40% of the target covered at 40X. Sequencing was performed at the McDonnell Genome Institute at Washington University School of Medicine in St. Louis.

Additionally, we performed low-pass whole genome sequencing for the longitudinal study as previously described (25). The resulting BAM files were used as the input data for the CNA characterization pipeline.

### SNV characterization

We used our previously reported software ITHE (20) to calculate by-patient SNV burden and divergence, leveraging the two neoplastic geographically distant samples and a control sample from the same patient. We recently developed, optimized, and validated this pipeline using 28 pairs of technical replicates (same extracted DNA, two aliquots were independently sequenced) of macrodissected FFPE DCIS samples similar to the specimens analyzed here. We used the filtering parameters we found optimal previously (20). ITHE was optimized for accurate divergence estimation and thus tries to maximize variant calling’s precision. We measured SNV divergence as the percentage of mutations detected in the union of the mutations from the two samples that are not shared by both samples. We required that the union set of mutations had at least five mutations to calculate divergence. SNV burden was calculated as the union of mutations in both samples. When comparing DCIS and IDC samples in the cross-sectional study, we report the mean of the two comparisons between one of the two DCIS samples and the IDC sample.

### Functional analysis

We performed the functional enrichment analysis of genes that harbored non-synonymous SNV mutations with PANTHER (26) and DAVID (27,28). We corrected the fold enrichment p-values considering the false discovery rate (FDR).

### CNA characterization

We followed our previously published protocols for low-pass WGS data processing and CNA calling (25). Briefly, we used Nextflow-base’s Sarek pipeline to align the lpWGS data to the GRCh38/hg38 reference genome, marked duplicates, and re-calibrated quality scores. We used the resulting alignments to call autosomal CNA variants using QDNAseq (29) on 50-kb genomic bins after filtering genomic regions and reads for mappability and QC content while estimating ploidy and purity. We corrected the log2 ratio for the latter. CNAs with |*corrected log*2 *radio*| > 0.3 were considered as altered and normal otherwise. To maximize the robustness of our statistics, we measured CNA burden per sample as the proportion of the genome that was altered (over the total genome considered) and CNA divergence per patient as the proportion of the altered genome that is not shared between the two samples over the altered genome per patient (i.e., *CNA divergence* = 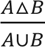, with *A* and *B* defined as the set of altered genomic regions of each homonymous sample, and Δ the set symmetric difference operator).

### Immunohistochemistry characterization

We chose a series of 15 candidate proteins (Supplementary Table S1) representing several categories including essential breast cancer drivers (ER, PR, HER2), immune-related (FOXP3, CD68), resource and microenvironmental measures (GLUT1, CA9, CD31, FASN), myoepithelial and basement membrane (TP63, COL15A1) and progenitor or stem cell-related (ALDH1 and RANK) markers. Additional proteins included the proliferation marker KI-67, the adhesion marker phospho-FAK, and COX2 (PTGS2), which were previously described as being associated with DCIS progression. In the longitudinal study, these were reduced to ER and GLUT1 only (Supplementary Tables S2-3), based on the results from the cross-sectional study and the paucity of samples. We measured stain intensity using detailed expert scoring. In most cases, the study pathologist used a scoring system that captures the distribution of intensities in an IHC profile, while for a smaller number of markers, it was binary (Supplementary Table S1). The IHC profile was quantified as the percentage of the slide presenting different levels of increasing staining intensity: absence, low, medium, and high. Medium staining was deemed as approximately twice as intense as low staining and high staining three times as intense as low staining.

We evaluated the IHC at three different scales of comparison:

1. The average intensity of immunofluorescence across samples for each patient, measuring the typical intensity of IHC signal per patient.
2. The variance of the intensity between samples for each patient, measuring the variations of IHC signal between distant locations in each patient.
3. The variance of intensity within samples, measuring the variations of IHC signal at short distances in each patient.

These three measures are quantified by the Mean of Intensity Score, the Earth Mover’s Distance, and the Cumulative Density Index. Briefly, the Intensity Score is the weighted sum of the IHC profile proportions normalized by the maximum possible staining, the Earth Mover’s Distance represents the minimum cost of turning one profile into another (30), and the Cumulative Density Index represents how close from a uniform distribution the observed profile is and ranges from 0 (all the profile weight in one of the extreme categories) to 1 (uniform profile). See a detailed description of these statistics in Supplementary Text 1.

### Statistical analysis

#### Cohort characterization

For each study, we compared differences in the central tendency of genetic and phenotypic variables per patient between cohorts using the Kruskal-Wallis Rank Sum or the Mann-Whitney U tests for many or two cohorts, respectively. We followed the Kruskal-Wallis Rank Sum test with Dunn’s post-hoc test while controlling for multiple tests using the Holm-Šidák adjustment (31). Exceptionally, CNA divergence met the assumptions of a parametric test, and thus, we used an ANOVA followed by Tukey HSD post-hoc tests. In cases where we used multiple measurements per patient (CNA burden), we used a Mixed-effects ANOVA with different random effect intercepts per patient to account for data dependencies (on the square-root-transformed variable), followed by Tukey’s HSD on the estimated marginal means.

#### Distinguishing Pure DCIS from Synchronous DCIS

We performed variable selection among the phenotypic measurements with significant differences between cohorts using a Random Forest classification model (32) under the Gini impurity criterion to return the importance ranking of each feature given by their predictive power. We used the two top measurements to build a generalized linear logistic model. Similarly, we built a generalized linear logistic model with the genetic measurements that showed significant differences between cohorts and the combination of the three. Due to missing data, we compared the models under the Akaike information criterion (AIC) on the smallest dataset for all models (33).

#### Association with clinical outcomes

Using our longitudinal study, we determined whether genetic and phenotypic statistics were independently associated with the time to clinical outcome (non-invasive recurrence or progression) using Cox regression analyses after checking they met the proportional hazards assumption. *Nonrecurrent* patients were right-censored using their follow-up time, and *progressors’* recurrence time was used as their time to clinical outcome. *Recurrents* were discarded when considering progression, and *progressors* were discarded when considering non-invasive recurrence. We also provide supplementary results in which the clinical outcomes are “any recurrence” and “progression without discarding *recurrent* patients.” In this case, *recurrents* were right-censored at the time of recurrence when considering progression, and otherwise, their recurrence time was used as their time to clinical outcome. We evaluated the statistical significance of Cox regressors using the Wald test. We used the proportional hazard regression model for one variable (SNV burden) to stratify patients into low and high SNV burden and plotted their event-free survival curves. We stratified using the risk relative to the patient with all variables (i.e., SNV burden here) set at the mean value (i.e., type = “risk”, reference = “sample”, in the *predict.coxph* function of the *survival* R package). We chose the threshold that maximized Youden’s J statistic (34) using the true outcomes. In all cases, we used the log-rank test to compare the survival trends of two or more groups.

We also integrated 18 clinical covariates (Supplementary Table S4) with our eight genetic and phenotypic measurements to model time to non-invasive recurrence and time to progression. We performed variable selection using Cox LASSO and chose the regularization parameter that minimized the partial-likelihood deviance via 10-fold cross-validation. To reduce the stochasticity of the results, we performed this process 100 independent times per model and selected the variables that were selected in at least 90% of them. To reduce missingness, we performed mean imputation on the clinical covariates before variable selection. The selected variables were used to build the final Cox regression models using all patients with available (imputed) data for those variables. Alternatively, we selected patients with data for all covariates chosen without imputation. We used the final models to stratify patients as in the univariate proportional hazards regression above. In all cases, the model used to stratify patients and plot their event-free survival curves includes all the variables included in the forest plot. All variables were standardized to make hazard ratios (HRs) comparable, and thus, HRs are relative to a change of 1 standard deviation unless specified otherwise.

## Data Availability

All the sequencing data used in this manuscript is publicly available. The cross-sectional WES data at SRA with IDs (SRP298346 and XX) and the longitudinal WGS and WES data at HTAN dbGaP’s study accession phs002371.v6.p1.

### Reproducibility

Scripts to reproduce most data pre-processing and statistical analysis can be found at https://github.com/adamallo/ManuscriptScripts_DCISRecurrenceVsProgression.

## Results

### Study cohorts

We investigated DCIS progression to invasive cancer using two independent observational studies with different patients: a cross-sectional study and a longitudinal study (Fig. 1, Table 1). In the cross-sectional study (Fig. 1A), we compared DCIS samples from patients with DCIS only (*Pure DCIS,* n = 58) versus DCIS samples from patients with synchronous DCIS with invasive ductal carcinoma (*Synchronous DCIS*, n = 61). In the separate longitudinal study (Fig. 1B), we compared pure DCIS samples from patients who were treated and had long-term follow-up (median = 117 months, 95% CI [104, 132]). This cohort consisted of patients who progressed to IDC (*progressors*) (n = 56), patients who had a DCIS-only recurrence (*recurrents,* n = 69), or patients who did not recur during the follow-up interval (*nonrecurrents,* n = 99). In both studies, we characterized the genotype and phenotype of two formalin-fixed paraffin-embedded DCIS samples per patient, enabling measures of evolutionary divergence (see Methods). We also obtained a single sample of their IDC recurrence for some progressors.

### Cross-sectional study

#### Single Nucleotide Mutational Burden

*Pure DCIS* carried fewer SNVs per patient (mean 7.5 ± 10.6 standard deviation) than *synchronous DCIS* (10.4 ± 15.3), but this difference was not statistically significant (Fig. 2A).

**Figure 2.**
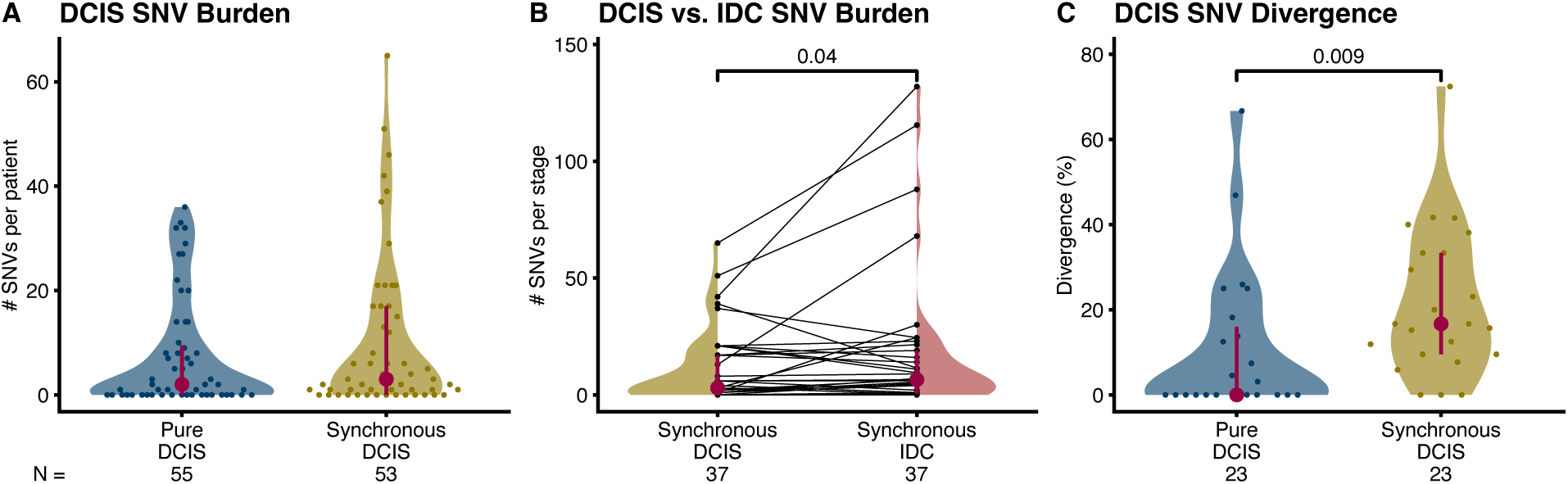
Cross-sectional SNV burden and divergence. Distribution of the number of SNVs per patient in the two cross-sectional cohorts **A** and the two lesion types (DCIS vs. IDC) present in the *synchronous* cohort **B.** Distribution of SNV genetic divergence (percentage of private mutations) per patient in the two cross-sectional cohorts **C.** We calculated divergence for tumors with at least five mutations in the union of the two samples, which explains the lower number of tumors per group. P-values shown if *p* ≤ 0.1, **A, C**: Mann-Whitney U, **B**: Paired-samples sign test. Interquartile range (vertical line) and median (point) in burgundy, N: number of patients.

The invasive component in *synchronous DCIS* patients showed a statistically significantly increased number of SNVs (18.1 ± 31.5, Fig. 2B) compared with their DCIS counterpart (Paired-samples sign test, *p* = 0.04) largely due to four cases of IDC with a dramatic increase in mutation burden.

#### SNV Genetic Divergence

We measured the SNV genetic divergence as the percentage of mutations that are private to either sample per patient. *Synchronous DCIS* showed higher genetic divergence (21.5% ± 17.5%) than *pure DCIS* (10.8% ± 17.4%, Fig. 2C) (Mann-Whitney U test, *p* = 0.009). Additionally, we also characterized the genetic divergence between the two synchronous components (i.e., DCIS vs. IDC in *synchronous* patients) (44.5% ± 29.0%), which is higher than the paired synchronous DCIS divergence (Supplementary Fig. S1, Paired-samples sign test, *p* = 0.002).

#### Phenotypic characterization

*Synchronous DCIS* samples presented higher levels of GLUT1 staining (*p* = 0.004) and lower levels of CA9 staining (*p* = 0.01) than *pure DCIS* samples (Fig. 3A, pairwise Mann-Whitney U tests of mean intensity scores [MIS], unadjusted p-values); all other markers showed non-significant differences between groups. This result holds when one of the two DCIS samples per patient is used randomly instead of the MIS (Supplementary Fig. S2).

**Figure 3.**
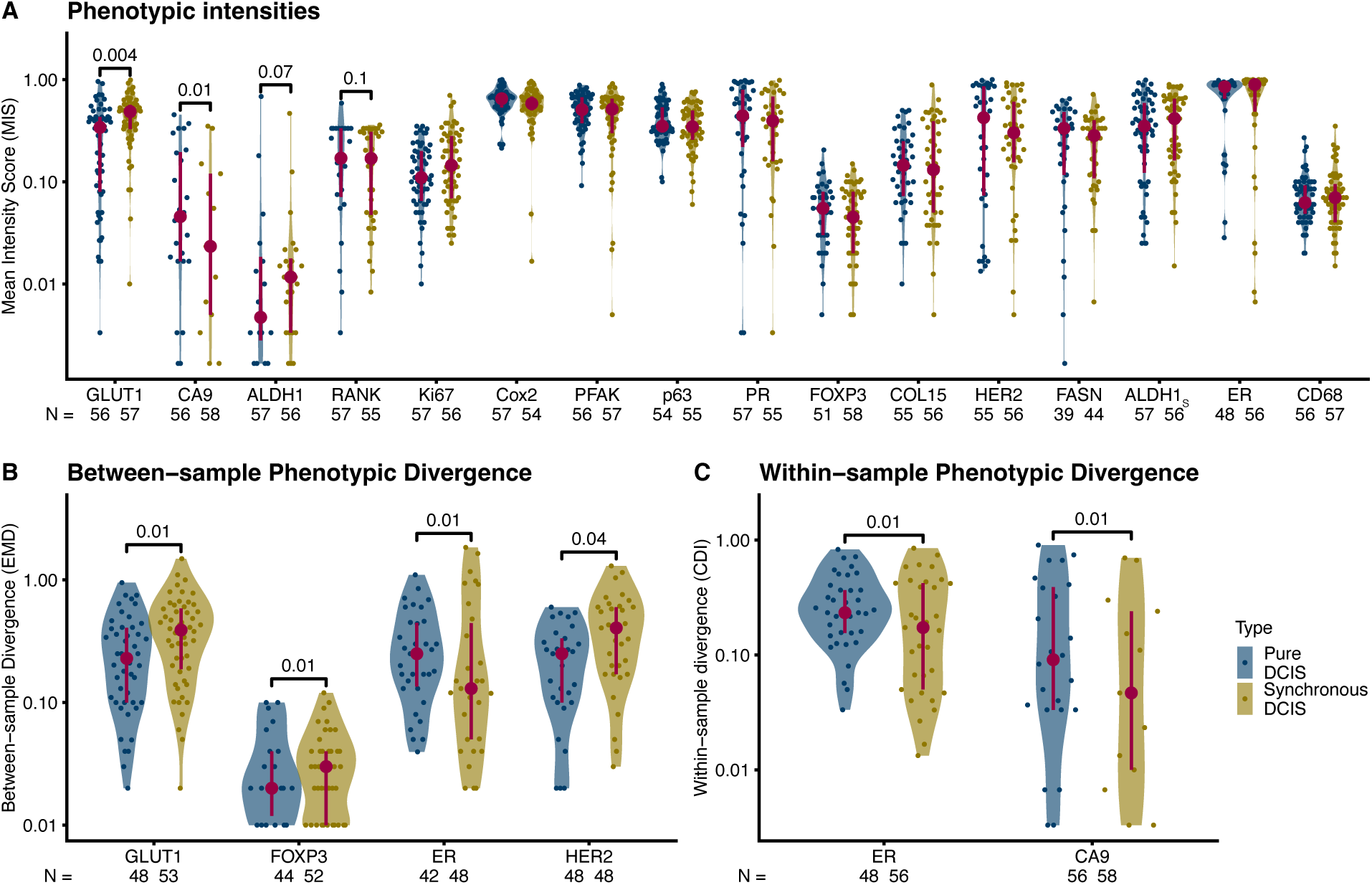
Cross-sectional phenotypic characterization and divergence. Distribution of mean intensity scores (MIS) per patient (see Methods) **A**, between-sample divergence (**B**, Earth Mover’s Distance [EMD]) and within-sample divergence (**C**, Cumulative Density Index [CDI]). **A:** for each patient and IHC marker, **B** and **C:** only markers with significant differences between cohorts (unadjusted p-values). Unadjusted pairwise Mann-Whitney U p-values shown if *p* ≤ 0.1. Interquartile range (vertical line) and median (point) in burgundy. N: number of patients.

#### Phenotypic Divergence

We characterized the between-sample phenotypic divergence for each marker using a distance between staining intensity profiles (Earth Mover’s Distance) and the within-sample divergence using a measure of staining intensity uniformity (Cumulative Density Index; see Supplementary Methods for detailed definition of these indices).

Multiple markers presented differences in between-sample divergence between *pure DCIS* and *synchronous DCIS* samples, with the latter showing increased divergence for GLUT1 (*p* = 0.01), FOXP3 (*p* = 0.01), and HER2 (*p* = 0.04) staining, but decreased divergence of ER (*p* = 0.01) staining (Fig. 3B, Supplementary Fig. S3, Pairwise Mann-Whitney U tests, unadjusted p-values). This reduction of ER phenotypic divergence in *synchronous DCIS* samples was replicated in the within-sample measures (*p* = 0.01) and mimicked by CA9 (*p* = 0.01) (Fig. 3C, Supplementary Fig. S4, Pairwise Mann-Whitney U tests, unadjusted p-values). A reduction in the phenotypic divergence for ER in *synchronous DCIS* samples indicates larger uniformity across and within samples, while the mean intensity of ER signal is not markedly different (Fig. 3A).

#### Distinguishing Pure DCIS from Synchronous DCIS

All eight significant phenotypic divergence features—MIS for GLUT1 and CA9 (Fig. 3A), EMD for GLUT1, FOXP3, ER and HER2 (Fig. 3B), and CDI for ER and CA9 (Fig. 3C)—were combined in a mixed logistic regression to model the progression status of the samples, from which the most important features were selected according to their relative predictive power. A reduced logistic model including between-sample diversity (EMD) for GLUT1 and within-sample diversity (CID) for ER had statistically significant coefficients (GLUT1 EMD, *p* = 0.01; ER CDI, *p* = 0.01) and spanned 40 pure DCIS cases and 52 synchronous DCIS cases. Therefore, we selected these two IHC markers (GLUT1 and ER) as the targets for phenotypic divergence to be included in the longitudinal study.

Logistic regression showed that the only statistically significant genetic measurement (SNV divergence) was strongly associated with the cohort, with *p* = 0.0136, so it was also selected for evaluation in the longitudinal study.

### Longitudinal Study: Associations with Recurrence and Progression

We used the cross-sectional cohort as a discovery cohort, using the synchronous DCIS as a proxy for high-risk DCIS likely to progress to IDC. Samples in our validation cohorts come from patients with pure DCIS with known outcomes (*nonrecurrent*, *recurred* as DCIS, *progressed* to IDC) and were obtained before treatment (Fig. 1B). We sequenced the exomes of two regions of each index DCIS in the longitudinal cohorts, mirroring the methods for the cross-sectional cohorts, and also performed low-pass whole genome sequencing data for most samples.

#### Mutational burden

Primary DCIS tissue from *nonrecurrent* patients carried the fewest SNVs (13.4 ± 18.2), followed by that of *recurrent* patients (19.2 ± 26.4) and *progressors* (39.7 ± 46.2). These relationships between cohorts were mirrored by the CNA alteration burden (*nonrecurrents*: 15.9% ± 15.0% genome altered, *recurrents*: 17.3% ± 14.8%, *progressors*: 24.6% ± 17.1%) but presented higher p-values. Thus, SNV burden shows statistically significant differences between *nonrecurrents* and *progressors* (*p* = 0.003) and between *recurrents* and *progressors* (*p* = 0.05, Dunn’s test corrected for multiple tests with the Holm-Šidák adjustment) (Fig. 4A). In contrast, CNA burden was significantly different only between *nonrecurrents* and *progressors* (*p* = 0.03, Tukey HSD) (Fig. 4B).

**Figure 4.**
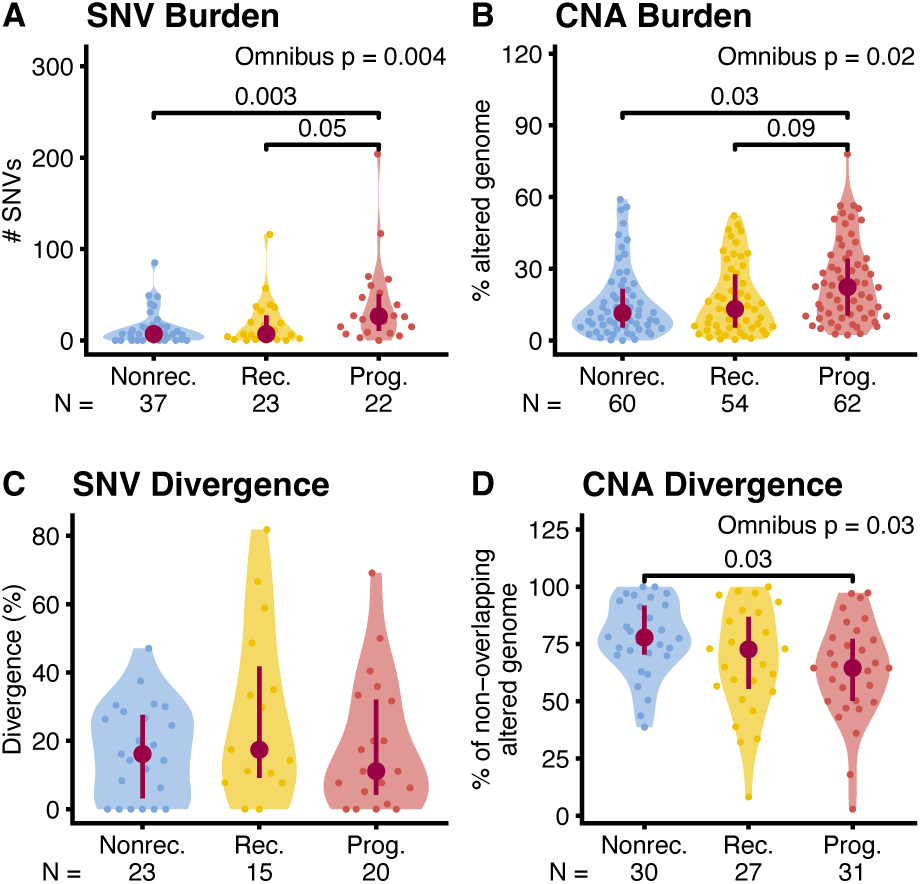
Longitudinal mutational burden and divergence. Distribution of SNV (**A**, **C**) and CNA (**B**, **D**) mutational burdens (**A**, **B**) and divergences (**C**, **D**) in the three longitudinal cohorts (Nonrec: *nonrecurrents*, Rec: *recurrents*, Prog: *progressors*). **A**: number of SNVs per patient; Omnibus test: Kruskal-Wallis Rank Sum, Post-hoc test: Dunn’s test with control for multiple tests using the Holm-Šidák adjustment. **B**: proportion of genome with copy number alterations per sample; Omnibus test: Mixed-effects ANOVA on the square-root-transformed proportion of genome altered, Post-hoc test: Tukey HSD on estimated marginal means. **C**: percentage of private SNV mutations per patient; Omnibus test: Kruskal-Wallis Rank Sum. **D**: percentage of the genome with copy number alterations private to either sample per patient; Omnibus test: ANOVA, Post-hoc test: Tukey HSD. P-values shown if adjusted *p* ≤ 0.1. Interquartile range (vertical line) and median (point) in burgundy, N: number of data points (**A, C,** and **D**: patients, **B**: samples). We only calculated divergence for tumors with at least five mutations in the union of the two samples, which explains the lower number of tumors in **C**.

#### Genetic Divergence

Similar to SNV divergence, we measured CNA divergence as the percentage of the altered genome that is private to either sample per patient. SNV divergence was highest in *recurrent* patients but not statistically different between cohorts (*nonrecurrents*: 17.0% ± 13.8%, *recurrents*: 28.2% ± 25.5%, *progressors*: 18.4% ± 19.3%, Fig. 4C). In contrast, CNA divergence followed a decreasing pattern of divergence with progression (Fig. 4D), by which *nonrecurrents* were the most divergent (77.4% ± 16.4%), followed by *recurrents* (67.7% ± 23.4%) and *progressors* (63.7% ± 21.7%). Only *progressors* and *nonrecurrents* showed statistically significant differences in CNA divergence in pairwise comparisons (Fig. 7B, p = 0.03, Tukey HSD).

#### Functional analysis of non-synonymous SNV mutations

The functional analyses highlighted significant differences between the three cohorts. According to *DAVID*, *recurrent* patients showed enrichment of mutated genes involved in taste reception (TAS2R30, TAS2R31, TAS2R43, and TAS2R46), while *progressors* showed enrichment of genes typically mutated in cancers such as endometrial, small cell lung, prostate, and breast cancer, glioma and melanoma (PIK3CA, ERBB2, PTEN, AKT1, PIK3R2, TP53, PIK3CG), and genes involved in the determination of cell shape, arrangement of transmembrane proteins, and organization of organelles (SPTA1, SPTBN5, DST, SPTAN1). *Nonrecurrents* did not show significant functional enrichment (Supplementary Table S5). In addition, *PANTHER* functional analysis revealed an enrichment of several pathways only in *progressors* (Supplementary Table S6), such as *Hypoxia response via HIF activation* (*p* < 0.001, false discovery rate correction herein this section), *Insulin/IGF pathway-protein kinase B signaling cascade* (*p* < 0.001), *p53 pathway* (*p* = 0.003), *Endothelin signaling pathway* (*p* = 0.003), *Hedgehog signaling pathway* (*p* = 0.02), and *PI3 kinase pathway* (*p* = 0.03).

#### Phenotypic Characterization and Divergence

We characterized the DCIS phenotypes of the three cohorts using the immunohistochemical profiles of the two markers that showed the highest discriminating power between the two cross-sectional cohorts, ER and GLUT1 (within-sample and between-sample divergence, respectively; see Immunohistochemistry characterization methods section). GLUT1 intensity was different between longitudinal cohorts (Fig. 5A, p = 0.04, Kruskal-Wallis Rank Sum), like in the cross-sectional study (Fig. 3A), with *progressors* having a generally higher intensity than nonprogressor cohorts, but the pairwise differences were not statistically significant (vs. *nonrecurrents p* = 0.06, vs. *recurrents p* = 0.07). ER intensity (Fig. 5B) was higher in ER+ *progressors* (*p* = 0.02) and *recurrents* (*p* = 0.03) than in *nonrecurrents* (Dunn’s test corrected for multiple tests with the Holm-Šidák adjustment). This new pattern was not found in the cross-sectional study, and the difference between *progressors* and *nonrecurrents* is robust to ER status stratification (Supplementary Fig. S5).

**Figure 5.**
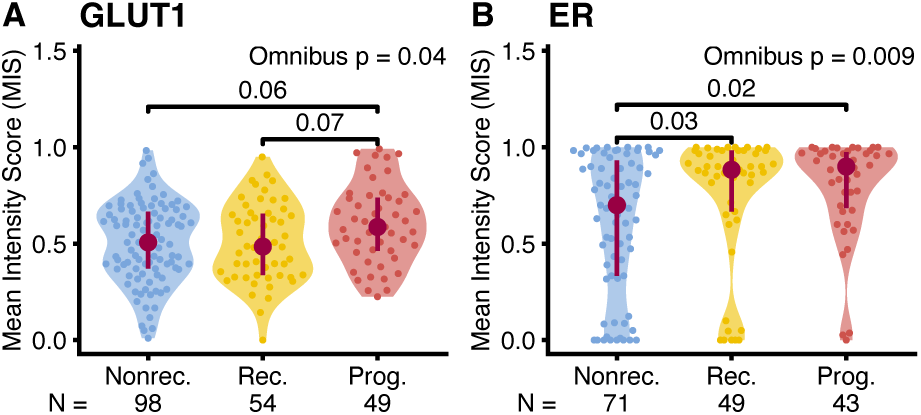
Longitudinal phenotypic characterization. Distribution of mean normalized intensities (MIS) per patient (see Methods) in the three longitudinal cohorts (Nonrec: *nonrecurrents*, Rec: *recurrents*, Prog: *progressors*). **A**: GLUT1 marker, **B**: ER marker in ER+ patients only. Omnibus test: Kruskal-Wallis Rank Sum, Post-hoc test: Dunn’s test with control for multiple tests using the Holm-Šidák adjustment. P-values shown if adjusted p ≤ 0.1. Interquartile range (vertical line) and median (point) in burgundy. N: number of patients.

We assessed the phenotypic divergence for these two markers using the same methodology as in the cross-sectional study, evaluating ER within-sample divergence and GLUT1 between-sample divergence, but neither showed a statistically significant difference between longitudinal cohorts (Supplementary Fig. S6).

#### Association with clinical outcomes

We tested if our genetic and phenotypic markers were independently associated with the time to non-invasive recurrence or progression using Cox regression analyses. Additionally, alternative clinical outcomes (recurrence [including progression] and progression with non-invasive recurrents right-censored) can be found in the supplementary materials (Supplementary Figs. S7-S8, S10, Supplementary Tables S7-S8).

Time to non-invasive recurrence was associated with divergences: SNV (*p* = 0.024), within-sample ER (*p* = 0.026), and CNA (*p* = 0.038), while time to progression was primarily associated with totals: SNV burden (*p* < 0.0001), ER intensity (*p* = 0.025), GLUT1 intensity (*p* = 0.027), and CNA burden (*p* = 0.045), but also CNA divergence (*p* = 0.025) (Supplementary Tables S9-S10, Wald test). The association between SNV burden and progression was the only one that survived multiple-test correction (Supplementary Tables S7-S8, progression adjusted *p* < 0.0001, Holm correction). Accordingly, we show the capability of this genetic measurement to stratify patients’ non-invasive-recurrence-free (Fig. 6A) and progression-free (Fig. 6B) survival by splitting patients into low and high SNV burden categories and comparing their event-free survival curves. The Kaplan-Meier plots show differences in the event-free survival curves, with median times to event that differ between groups 100 months for non-invasive recurrence (Fig. 6A, p = 0.026) and 57 months for progression (Fig 6B, *p* < 0.0001, Log-rank test).

**Figure 6.**
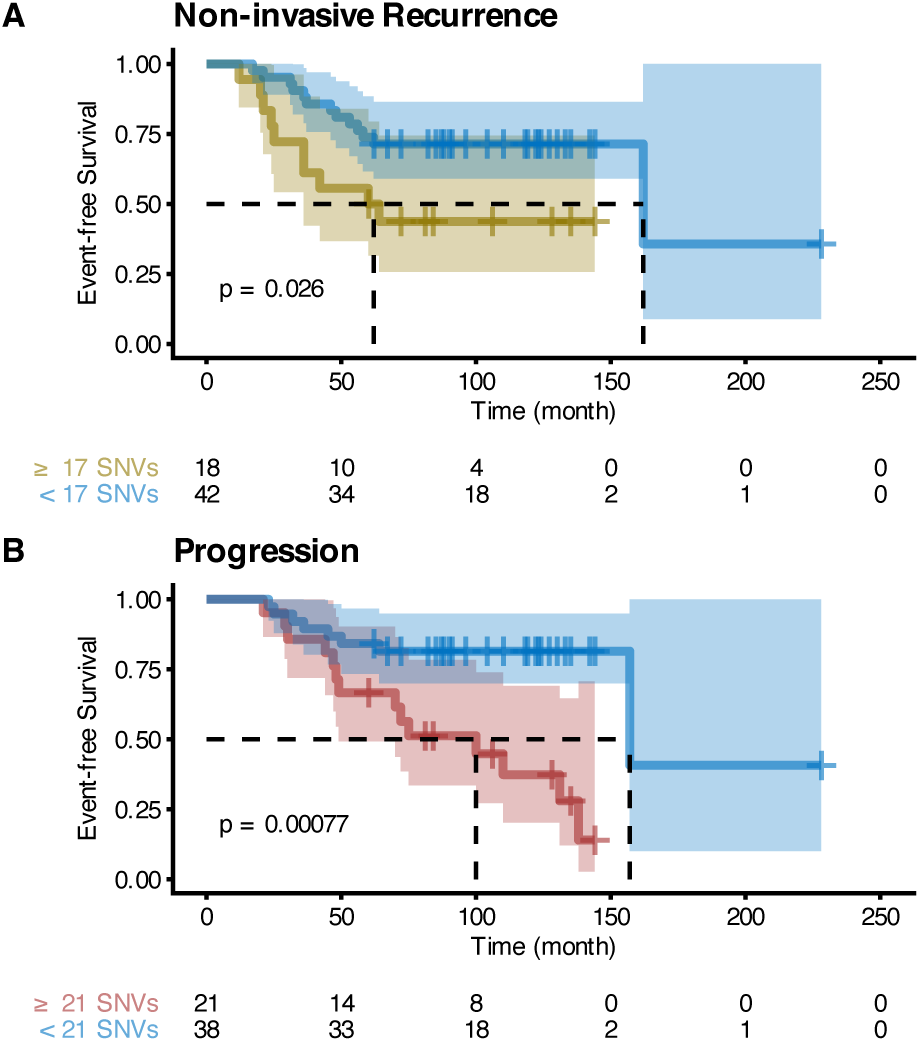
Event-free survival curves of patients stratified by SNV burden. Kaplan-Meier plots of stratified patients. **A**: Non-invasive-recurrence-free survival. **B**: Progression-free survival. SNV burden thresholds maximize Youden’s J statistic of the outcomes (17 SNVs for non-invasive recurrence and 21 for progression). Log-rank test. The table below the Kaplan-Meier plot shows the number of samples at risk at different times.

Finally, we integrated 18 clinical covariates (Supplementary Table S4) with our genetic and phenotypic measurements to develop comprehensive models of DCIS non-invasive recurrence and progression. Proportional hazard regressions built with variables selected using LASSO contained three significant variables for non-invasive recurrence (Fig. 7A, treatment option *p* < 0.001, ER status *p* = 0.003, and SNV divergence *p* = 0.018, Wald test) and two for progression (Fig. 7C, surgical margin *p* = 0.017, and SNV burden *p* = 0.004, Wald test), and event-free survival curves of patients stratified using their relative risk were highly significant, with median time to events that differ between groups in 123 months for non-invasive recurrence (Fig. 7B) and > 69 months for progression (Fig. 7D). An alternative parameterization of the surgical margin as a 2mm threshold showed very similar results (Supplementary Fig. S9, *p* = 0.048, Wald test). The associations with the treatment option and ER status were repeatable without using covariate imputation (Supplementary Fig. S10), while the surgical margin association was only robust when not excluding *recurrent* patients (Supplementary Figs. S11-S12). No other significant variables in these models were imputed.

**Figure 7.**
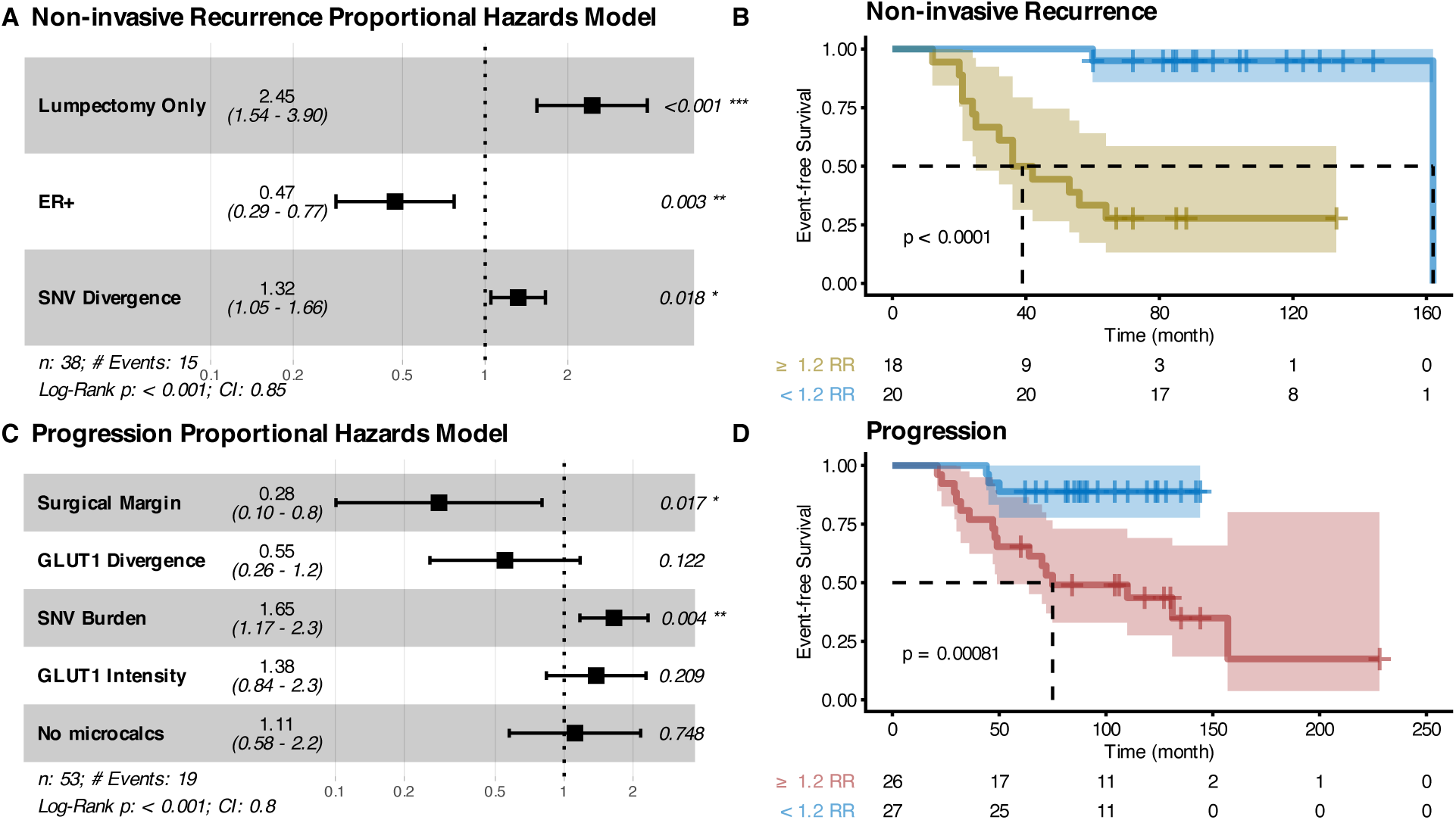
Associations with time to clinical outcome. Forest plots describing proportional hazard regressions using variables selected with LASSO (**A**, **C**) and corresponding Kaplan-Meier plots of patients stratified by the relative risk threshold that maximizes Youden’s J statistic of the outcomes (**B**, **D**). **A-B**: Non-invasive-recurrence-free survival. **C-D**: Progression-free survival. Hazard Ratios (second column, **A**, **C**) are relative to 1 standard deviation. Lumpectomy Only is compared to Lumpectomy + Radiation and Mastectomy and ER+ is compared to ER-. No microcalc(ification)s is compared to having microcalcifications in DCIS-only and/or benign ducts. Tables below Kaplan-Meier plots show the number of samples at risk at different times. Log-rank test.

## Discussion

Evolutionary measurements summarize the results of complex evolutionary dynamics, and equivalent observations may result from very different evolutionary scenarios. For example, both a low mutation rate under neutral evolution and a hard selective sweep can generate low divergence of high allele-frequency mutations. Divergence also has multiple scales, and multiple evolutionary processes may affect scales differently or even in opposite directions. Clonal expansion may reduce within-sample divergence but increase between-sample divergence. Intra-tumor heterogeneity provides the fuel for natural selection, but it is not clear what form of intra-tumor heterogeneity (genetic, epigenetic, or phenotypic) is most relevant to the clinical outcomes of a particular tumor, and it is not clear how best to measure it (18).

The improved efficacy of preventive screenings provided the ability to identify many tumors in the earliest phases of their evolution, demanding the development of new approaches to stratify the risk to these patients to avoid over- and undertreatment. However, every neoplasm develops a unique set of alterations through somatic evolution (18), making it unlikely that any given set of molecular markers will be universally applicable, even within a given cancer type. In contrast, measures of the evolvability of a neoplasm, such as the number of mutations and measures of intra-tumor heterogeneity, may be universal biomarkers that predict neoplastic progression in many different types of cancers and pre-cancers (9,10,15). By taking two spatially distinct samples for each primary pre-cancer, we measured genetic and phenotypic divergence within and between samples, and their relationship with two key clinical processes: 1) recurrence of precancer following treatment and 2) progression of precancer to invasive cancer.

### DCIS recurrence and progression are different biological processes

Based on our results, progression from DCIS to invasive breast cancer appears to be a qualitatively and biologically different process from recurrence of DCIS. We had assumed that progression to invasion first requires recurrence of the DCIS and so expected that the factors that predicted recurrence would also predict progression. We were surprised that there was no overlap in their multivariate models (Fig. 7).

Among all genetic and phenotypic variables, SNV burden, as measured with our previously released software ITHE (20), was the variable that showed the largest differences between the patients that did not recur, the patients that recurred with DCIS, and the patients that progressed to IDC. SNV burden also had the strongest independent association with time to progression and was an essential component of its best multivariate model. The lpWGS CNA burden from the same samples corroborated this finding with higher p-values. Theoretically, this increase in mutation burden may result from an increase in mutation rate, evolutionary time, or self-renewing cell population size. However, due to limitations in detecting variants at low allele frequency, measured mutation burdens are biased towards high allele frequency mutations and are thus most sensitive to early increases in mutation rates or the selective evolutionary forces that drive clonal expansion (35,36). This bias is especially true when using our program ITHE since, by design, it maximizes specificity in exchange for a lower sensitivity for low-frequency mutations in a sample. For these reasons, we do not necessarily expect SNV burden measured differently to show the associations found here.

Progression was also associated with two other magnitude measurements (i.e., totals: ER and GLUT1 intensities) but did not provide enough additional information over the SNV burden to be included as significant variables in the best multivariate model, which also included the size of the surgical margin as a significant predictor.

Previous studies have shown that surgical margins are clinically important in reducing the risk of ipsilateral breast tumor recurrence after breast-conserving surgery (37,38). Positive margins (i.e., DCIS at the edge of the resected tissue) clearly increase recurrence risk, but patients with positive margins were excluded from our study. Instead, we analyzed how the size of the negative margins associate with the clinical outcome. The evidence for this association is mixed in the literature (37,39), but current consensus guidelines consider margins >2mm adequate. Notably, these studies do not typically differentiate recurrence of DCIS from progression to invasive disease in their endpoints, as we did here. We found that the size of the surgical margins was one of the strongest predictors of progression but was not a statistically significant predictor of recurrence with DCIS, neither in the selected multivariate model nor in isolation. This negative result may be due to a type II error, but even if such an association exists, it is likely to be weaker than that observed for progression. We hypothesize that a micro-invasive phenotype could reduce the probability of obtaining large surgical margins, or a phenotype that makes DCIS cells more independent could allow small clusters of cells left over during surgical treatment to survive and further progress to invasive disease more readily. This finding highlights the importance of segregating non-invasive recurrence from progression and how associations with recurrence (of any kind) are primarily a composite of the associations with non-invasive recurrence and progression (Supplementary Fig. S8A). We confirmed our results using the consensus guideline >2mm threshold instead of treating surgical margins as a continuous variable, obtaining equivalent though weaker results. This observation shows prognostic information in the size of the surgical margin. The fact that all associations with progression held independently of whether we excluded recurrent patients or right-censored them at the time of DCIS recurrence (Supplementary Fig. S8C-D) shows their robustness and adds evidence towards non-invasive recurrence and progression being qualitatively different phenomena.

In contrast, time to non-invasive recurrence was associated with the extent of genetic divergence of SNVs between the two assayed regions of DCIS. We could not corroborate this finding with CNA divergence, which followed the opposite trend but was also correlated to time to recurrence in the univariate models. The true (i.e., known without error) amount of genetic divergence measured using different mutation types should yield equivalent results if large enough mutational burdens of both types are accumulated. A few estimation biases may explain the discordance we observed between SNV and CNA divergences. A low CNA burden may increase the estimated divergence due to a higher false positive rate in the segmentation process without a broad range of true relative intensity values. In fact, CNA burden and CNA divergence were moderately anticorrelated across the study (ρ = −0.36, p < 0.001), and this anticorrelation was driven by the cohort with the lowest CNA burden. High within-sample heterogeneity is also expected to reduce the accuracy of between-sample divergence estimates and lead to the underestimation of the mutation burden. Low SNV burden also leads to missing data in SNV divergence estimates since divergence cannot be calculated accurately with few alterations. ER divergence followed the same direction as CNA divergence, with greater divergence associated with a lower risk of recurrence, but SNV divergence followed the opposite trend. These divergences were the only three measurements associated with time to non-invasive recurrence in the univariate analyses (Supplementary Table S9). Non-invasive recurrence is associated exclusively with divergence statistics, while progression was primarily associated with totals (SNV burden and mean GLUT1 intensity). Intratumor heterogeneity can arise from an increase in the amount of evolution (same mechanisms as mutation burden above) but also with diversifying selection, and we have previously associated it with poor prognosis in other pre-cancers (9).

Non-invasive recurrence was also associated with the type of DCIS treatment and estrogen-negative status. The fact that patients treated with lumpectomy alone were more likely to recur than those treated with lumpectomy and radiation or mastectomy has been well described. Adjuvant radiation therapy has been previously shown to reduce the risk of recurrence (40), and after mastectomy, patients are no longer screened using mammograms, making it unlikely that asymptomatic noninvasive recurrences would be detected. The association between recurrence and ER status may be unsurprising since patients with ER+ breast cancers have better prognoses than ER-ones (41,42). However, its association with DCIS recurrence is unclear (43–45), and the balance of evidence points against it (46). As for surgical margins, most studies are limited by not differentiating between recurrence and progression endpoints. At least one of the studies that made this distinction (43) found a decrease in non-invasive, but not in invasive recurrences in ER+ patients, which is consistent with our results. Different endpoints may partially explain the mixed evidence on the association between ER and DCIS recurrence and progression.

Functional genetic analysis also showed a difference between the three cohorts, particularly between those DCIS that recurred compared to those that progressed. DCIS that will recur without invasion shows enrichment of mutations in genes involved in the TAS2R signaling network. The activation of these genes determines a pro-apoptotic, anti-proliferative, and anti-migratory response action in highly metastatic breast cancer cell lines (47). These genes also appear to be involved in the regulation of apoptosis in head and neck squamous cell carcinoma, and their impairment could favor the survival of cancer cells (48). On the other hand, DCIS that will progress to invasion demonstrates a broader variety of biological processes and pathways involved, such as hypoxia response, insulin/IGF, endothelin, hedgehog, p53, and PI3 kinase signaling pathways. These biological processes are typically altered in various types of cancer and also show an enrichment of mutations in genes involved in the reorganization of the cytoskeleton. The ability to metastasize outside the mammary gland and to relapse observed in these patients is supported by mutations in those pathways.

### Synchronous DCIS is not a good model for DCIS progression

Cross-sectional studies are much less resource-intensive, faster, and simpler to conduct than longitudinal cohort studies. If synchronous DCIS (adjacent to IDC) was a good model for primary DCIS that later progressed to IDC, cross-sectional studies could be more readily employed as relevant surrogates for cancer progression. However, our results show this is not possible for our purpose, and in fact, synchronous DCIS shares more similarities with DCIS that will recur as DCIS than with DCIS that will progress.

The pure DCIS samples in our cross-sectional study are equivalent to a mixture of samples from the three cohorts in our longitudinal study since their future outcomes are not considered. Thus, characteristics associated with clinical outcomes are expected to be mixed in the cross-sectional study. We found that DCIS adjacent to IDC showed increased divergence, which may result from divergent evolution facilitated by longer evolutionary times, the interaction with IDC, or an intrinsic characteristic of early-progression DCIS. If we assume that IDC originates from DCIS (stepwise progression model), synchronous DCIS samples are (on average) evolutionarily older than pure DCIS samples, representing a later evolutionary stage than samples from either study. In this case, the cross-sectional study would reveal differences between early and late DCIS. Alternatively, if we assume that an early progression model is also possible (i.e., born to be bad (49)), synchronous DCIS would be enriched with this DCIS sub-type. In this case, the cross-sectional study would show evolutionary characteristics that distinguish those DCIS fated for invasive progression. Additionally, the presence of IDC near synchronous DCIS may also alter its characteristics, modifying its environment systemically (e.g., immune response) and locally (e.g., microenvironment and cell composition through cell migration).

The higher between-sample genetic divergence we found in synchronous DCIS compared to pure DCIS aligns better with stepwise DCIS progression, in which late DCIS would have had more evolutionary time to undergo divergent evolution. Under the early progression model, this may be an intrinsic characteristic of such a DCIS subtype that could facilitate the rapid invasion of nearby tissues. Most (75%) markers with significantly different between-sample divergences showed higher divergence in synchronous DCIS, and all markers with significantly different within-sample divergences showed the opposite trend. These results are concordant with the genetic results and our expectations under a stepwise progression model but did not survive multiple-test correction.

### Integrating the results with clonal evolution in neoplastic progression

The two observational studies we conducted here are complementary and together improve our understanding of the evolutionary process leading to DCIS progression and recurrence. We find that primary DCIS that will progress to IDC is more genetically and phenotypically evolved, with higher SNV and CNA burden and more aggressive phenotypes, both metabolically and with respect to its estrogen sensitivity. At least one selective sweep is likely a part of their evolutionary history, which would reduce genetic divergence in the tumor. Higher cell motility could also reduce between-sample heterogeneity. Surgical margins show the strongest association with progression, suggesting that there may be features of the growth pattern of these lesions that make it more difficult to completely excise surgically. In contrast, DCIS recurrence may be primarily enabled by suboptimal clinical management. The few evolutionary features associated with DCIS recurrence suggest an increased accumulation of evolutionary changes in those lesions compared to those that do not recur, which nevertheless do not attain the degree of divergence necessary for invasive progression. In aggregate, the evolutionary history of DCIS recurrences may lack the strong selective sweeps that may be necessary conditions to invade other tissues successfully. DCIS adjacent to IDC shows increased divergence, which may result from divergent evolution facilitated by longer evolutionary times, the interaction with IDC, or an intrinsic characteristic of early-progression DCIS (i.e., born to be bad).

### Conclusions

In summary, the evolutionary and clinical measures that predict the recurrence of DCIS differ from those that predict progression to IDC. Furthermore, DCIS adjacent to concurrent invasive cancer appears to be distinct from DCIS that will progress to invasive cancer over time. These findings suggest that the biological dynamics that make DCIS likely to recur differ from those that make it likely to progress, and those dynamics interact differently with our clinical interventions. These insights have the potential to improve both risk stratification and individualized patient management for high-risk DCIS.

## Data Availability

All raw data will be publicly available at publication. Some of our raw datasets are still being uploaded to SRA, and placeholders in the draft have replaced their accession numbers. The cross-sectional WES data is available at SRA with IDs (SRP298346 and XX) and the longitudinal WGS and WES data at HTAN dbGaP's study accession phs002371.v6.p1.

## Acknowledgments

We thank the Research Computing at Arizona State University for providing HPC (50) and storage resources that have contributed to the research results reported here. This work is supported in part by NIH grants U54 CA217376, U2C CA233254, R21 CA257980, and R01 CA140657, as well as CDMRP Breast Cancer Research Program Award BC132057 and the Arizona Biomedical Research Commission grant ADHS18-198847. The findings, opinions, and recommendations expressed here are those of the authors and not necessarily those of the universities where the research was performed or the National Institutes of Health.

