## Supplementary Materials for "Evolutionary Measures Show that Recurrence of DCIS is Distinct from Progression to Breast Cancer"

#### Table of contents

**Supplementary Text 1: Immunohistochemistry Distances** **2**
- IHC measurements 2
- Mean of Intensity Score (MIS) 2
- Earth Mover's Distance (EMD) 2
- Cumulative Density Index (CDI) 3
- Controlling for triple-negative breast cancer 3
- References 4

**Supplementary Figures** **5**
- Figure S1. Cross-sectional SNV divergence within synchronous DCIS and between synchronous DCIS and IDC. 5
- Figure S2. Cross-sectional phenotypic characterization. 6
- Figure S3. Cross-sectional phenotypic between-sample diversity. 7
- Figure S4. Cross-sectional phenotypic within-sample diversity. 8
- Figure S5. ER longitudinal phenotypic characterization in all patients. 9
- Figure S6. Longitudinal phenotypic divergence. 10
- Figure S7. Event-free survival curves of patients stratified by SNV burden: alternative clinical outcomes. 11
- Figure S8. Associations with time to clinical outcome: alternative clinical outcomes. 12
- Figure S9. Associations with time to clinical outcome: alternative clinical margin covariate. 13
- Figure S10. Associations with time to clinical outcome without covariate imputation. 14
- Figure S11. Associations with time to clinical outcome without covariate imputation: alternative clinical outcomes. 15
- Figure S12. Associations with time to clinical outcome without covariate imputation: alternative clinical margin covariate. 16

**Supplementary Tables** **17**
- Table S1. Breakdown of the number of patients for each monoclonal immunohistochemical marker in the cross-sectional study. 17
- Table S2. Breakdown of the number of patients for each monoclonal immunohistochemical marker in the longitudinal study. 17
- Table S3. Breakdown of the number of patients for each monoclonal immunohistochemical marker in the longitudinal study restricted to ER+ patients. 18

Table S4. Clinical variables considered in the proportional hazard regressions of the longitudinal study. 18
Table S5. DAVID functional annotation clustering results. 19
Table S6. PANTHER overrepresentation test results using the Fisher test and FDR correction. 21
Table S7. Univariate proportional hazard regressions of Time to Recurrence. 22
Table S8. Univariate proportional hazard regressions of time to progression from nonprogressors 23
Table S9. Univariate proportional hazard regressions of time to non-invasive recurrence 23
Table S10. Univariate proportional hazard regressions of time to progression 24

### Supplementary Text 1: Immunohistochemistry Distances

#### IHC measurements

An IHC profile is a set of categories indicating the percentage of the slide presenting basic levels of staining intensity: {$C_0$, $C_1$, $C_2$, $C_3$}, where $C_0$ is the fraction of the block with no staining, and $C_1$, $C_2$, and $C_3$ are the fractions of the sample with low, medium and high staining intensity.

#### Mean of Intensity Score (MIS)

For an IHC marker profile, the total intensity score is defined as the weighted sum of intensity values normalized by the maximum possible staining (meaning the IHC profile where the whole slide has high staining intensity):

$$I = \left(\frac{1}{100n}\right)\sum_{i=0}^{n} i \cdot C_i$$

Where $C_i$ is the fraction of the block with staining level *i*. The Mean of Intensity Score is the mean across blocks for each patient, measuring the typical observed staining intensity for that marker.

#### Earth Mover's Distance (EMD)

We consider the differences in the IHC profile between blocks for each patient to assess (distant) phenotypic heterogeneity in the tissue. EMD is a Wasserstein metric representing the minimum cost of turning one profile into another (1). The general cost function is computed as the product of the fraction of the profile and the distance that fraction is moved (measured as the number of categorical steps required to turn one profile into the other). For instance:

1. The score between profiles {100, 0, 0 , 0} and {0,100,0,0} is
   **EMD**({100, 0, 0 , 0}, {0,100,0,0}) = 1.0 x 1 = 1,
   because all the units are moved one level up.

2. In the same way,
   **EMD**({100, 0, 0 , 0}, {50,0,50,0}) = 0.5 x 2 = 1
   because half the units are moved two levels.

3. And
   **EMD**({100, 0, 0 , 0}, {0,0,0,100}) = 1.0x3 = 3,
   which is the maximum possible distance for this metric.

EMD is a pairwise metric that returns a distance measure, defined between zero, for identical profiles and (n-1) for maximally dissimilar profiles with n levels. We used the package *emdist* (version 0.3-2) implemented in R to evaluate this metric.

#### Cumulative Density Index (CDI)

We consider a measure of within-sample heterogeneity defined as follows:

$$\text{CDI} = 1 - \left|\frac{\sum_{i=0}^{n}(S_i - L_i)}{\sum_{i=0}^{n}(1 - L_i)}\right|$$

with $S_i = \sum_{i=0}^{i} P_j$ the cumulative values of the normalized profile, $L_i = \frac{i+1}{n+1}$ the cumulative values of a uniform profile (i.e., {0.25, 0.25, 0,25, 0.25} giving a cumulative {0.25, 0.50, 0.75, 1.0}). The denominator term is the same as the area under the cumulative uniform profile, and the numerator term is the area between the observed cumulative profile and the uniform one. With this definition, CDI represents how close to a uniform distribution the observed profile is. If the observed profile is uniform, then CDI=1, and if the observed profile is one of the extremes (e.g., {100, 0, 0, 0} or {0,0,0,100}), then CDI=0 (see Figure ST1).

This score is particularly sensitive to distinguishing extreme staining cases (i.e., all-or-nothing cases) versus cases with more diverse profiles. Values of CDI close to 0 indicate **less heterogeneity**, while values close to 1 signify more diverse profiles. A uniform profile, CDI=1, would indicate an equal representation of staining intensity in all levels, which is the maximum level of heterogeneity possible in the sample.

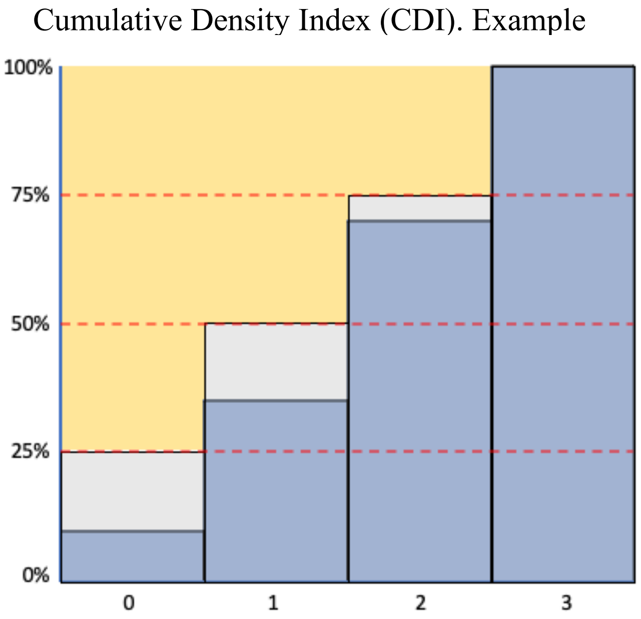


**Fig ST1:** Grey boxes represent the difference between the observed cumulative profile (blue) and a reference uniform cumulative profile ({0.25, 0.50, 0.75, 1.00}). The yellow area represents the area under the uniform profile. The value of CDI is given by the ratio between the grey area and the yellow area.

#### Controlling for triple-negative breast cancer

To control for the possible confounding effect of triple-negative breast cancer samples, which are known to have an elevated risk of subsequent breast events, we did an additional analysis in which we eliminated cases with negative ER status (PR and HER2 status were not known for many cases) for all of the regular IHC staining, additional standard clinical IHC, and RNA. Their phenotypic characterization and diversity were not qualitatively different, and their clinical outcome prediction results were very similar.

### Supplementary Figures

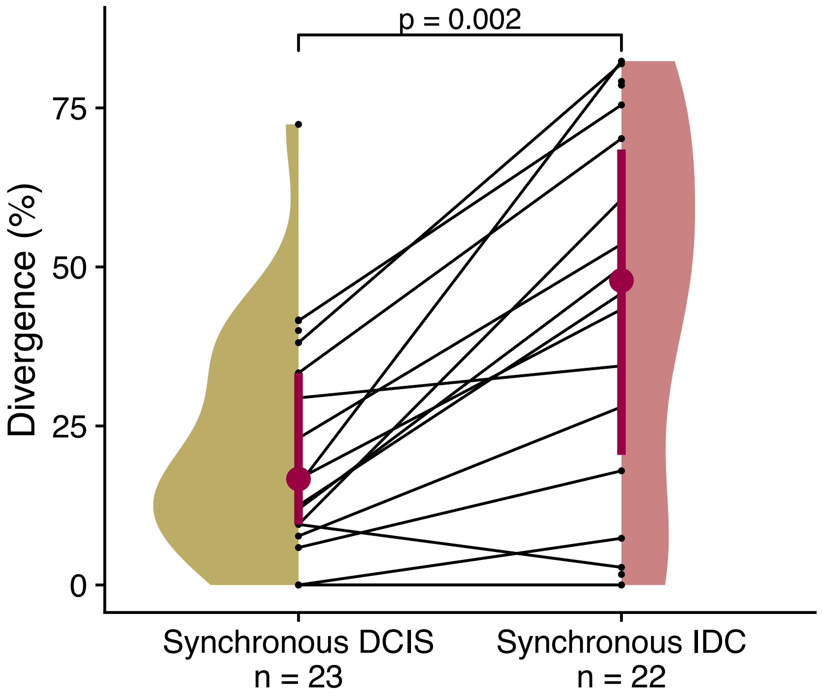


**Figure S1. Cross-sectional SNV divergence within synchronous DCIS and between synchronous DCIS and IDC.**
Distribution of SNV genetic divergence (percentage of private mutations) per synchronous patient, calculated within DCIS samples (Synchronous DCIS) or between DCIS and IDC samples (Synchronous IDC). Synchronous IDC data points are the mean of two comparisons between 2 DCIS samples and 1 IDC sample. Paired-samples sign test. Interquartile range (vertical line) and median (point) in burgundy. N: number of data points.

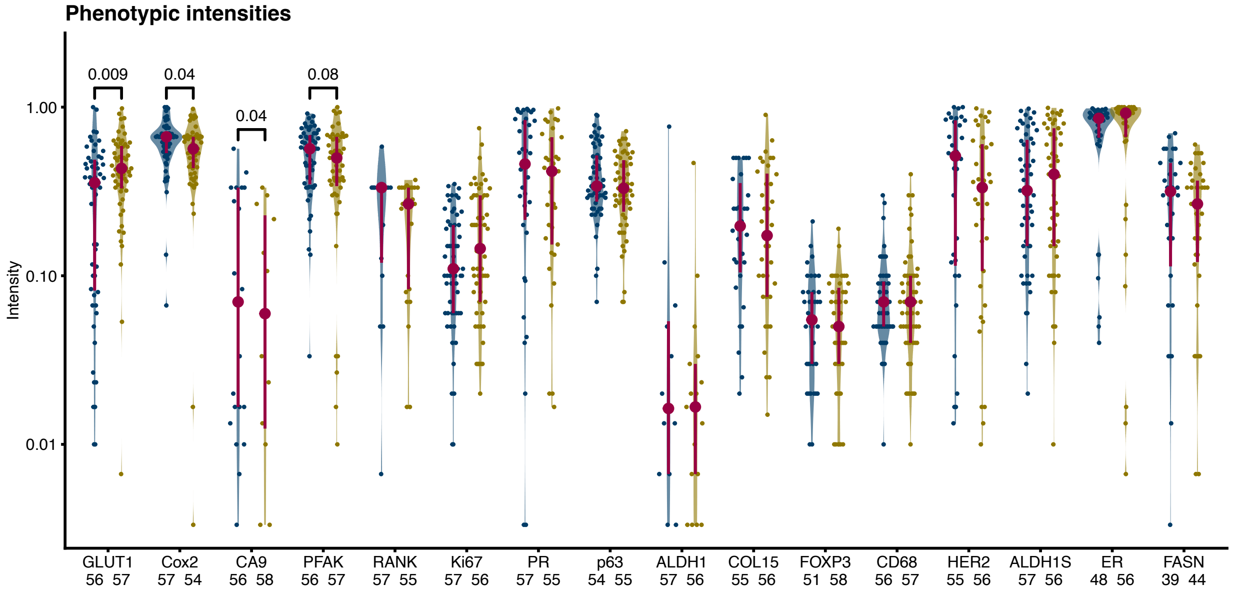


**Figure S2. Cross-sectional phenotypic characterization.**

Distribution of intensity scores of one sample per patient sampled at random for each patient and IHC marker (unadjusted p-values). Unadjusted pairwise Mann-Whitney U p-values shown if $p \leq 0.1$. Interquartile range (vertical line) and median (point) in burgundy. N: number of patients.

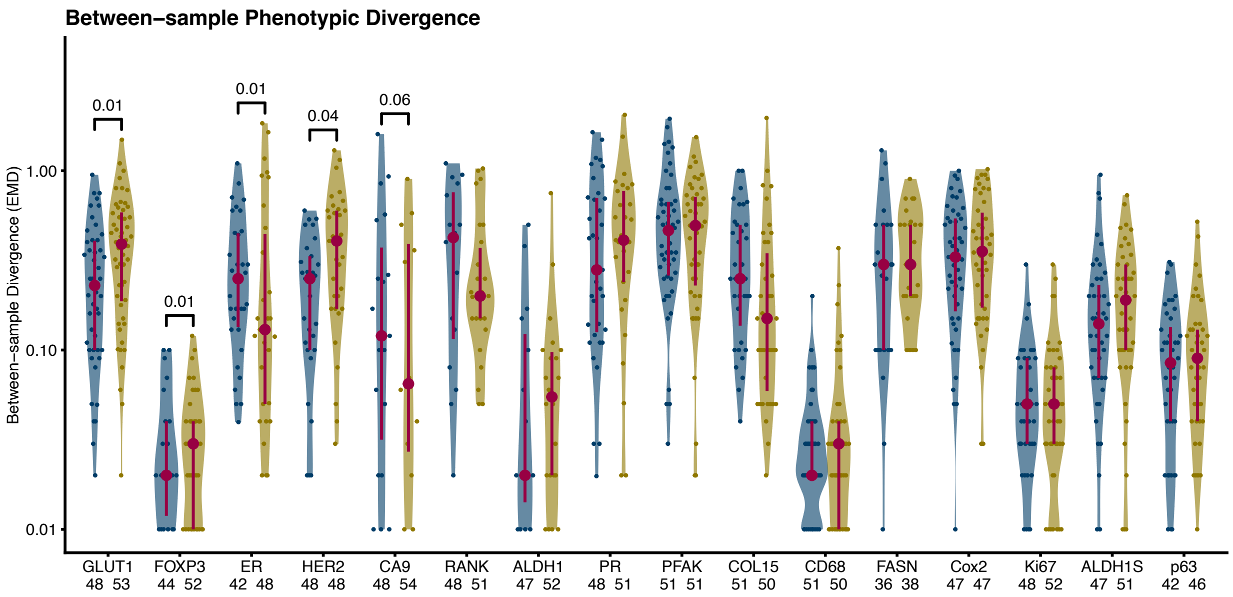


**Figure S3. Cross-sectional phenotypic between-sample diversity.**
Distribution of Earth Mover's Distances (EMDs) for each patient and IHC marker. Unadjusted pairwise Mann-Whitney U p-values shown if $p \leq 0.1$. Interquartile range (vertical line) and median (point) in burgundy. N: number of patients.

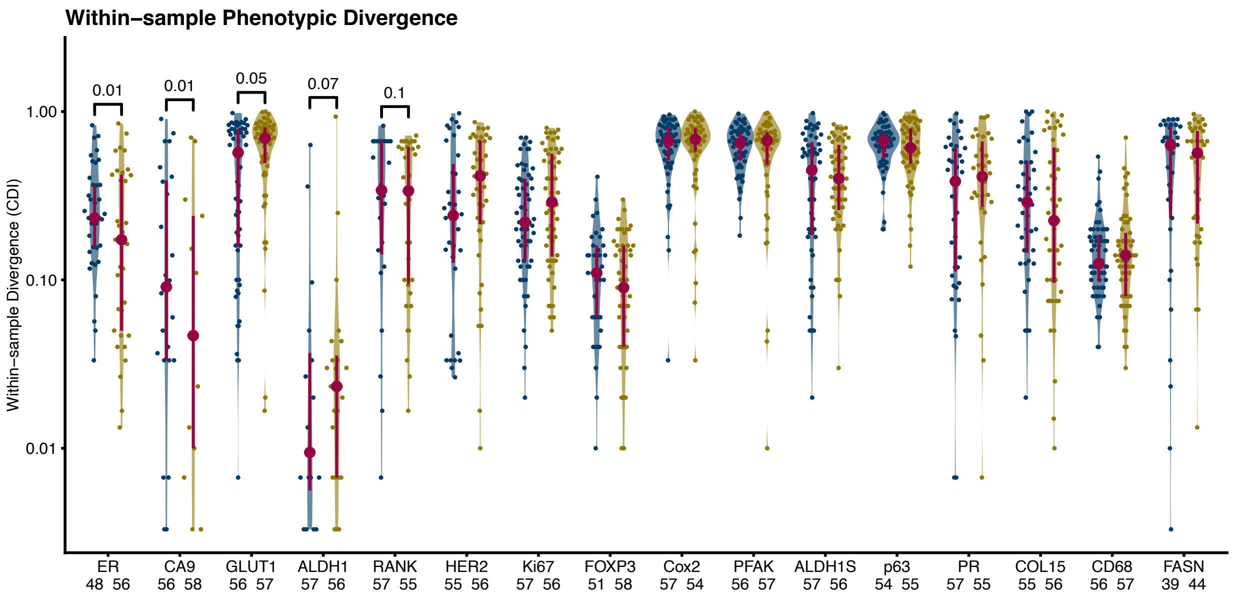


**Figure S4. Cross-sectional phenotypic within-sample diversity.**
Distribution of Cumulative Density Indices (CDIs) for each patient and IHC marker. Unadjusted pairwise Mann-Whitney U p-values shown if $p \leq 0.1$. Interquartile range (vertical line) and median (point) in burgundy. N: number of patients.

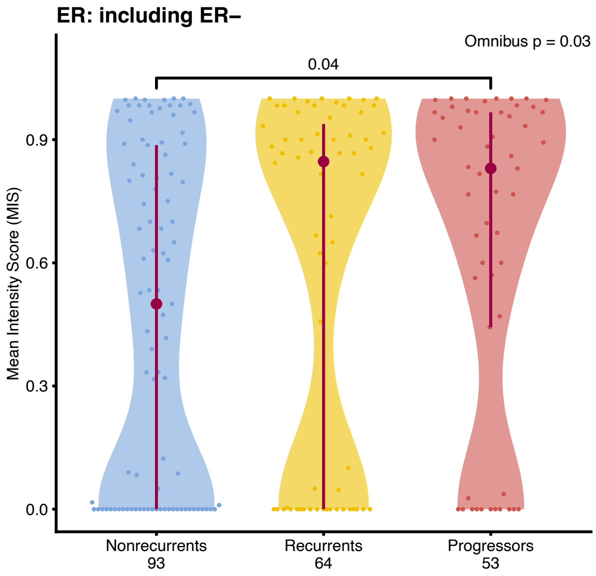


**Figure S5. ER longitudinal phenotypic characterization in all patients.**
In contrast with Fig 5B, ER- patients are included here. Distribution of mean normalized intensities (MIS) per patient (see Methods). Omnibus test: Kruskal-Wallis Rank Sum, Post-hoc test: Dunn's test with control for multiple tests using the Holm-Šidák adjustment. Statistically significant differences between groups shown if adjusted $p \leq 0.05$. Interquartile range (vertical line) and median (point) in burgundy. N: number of patients.

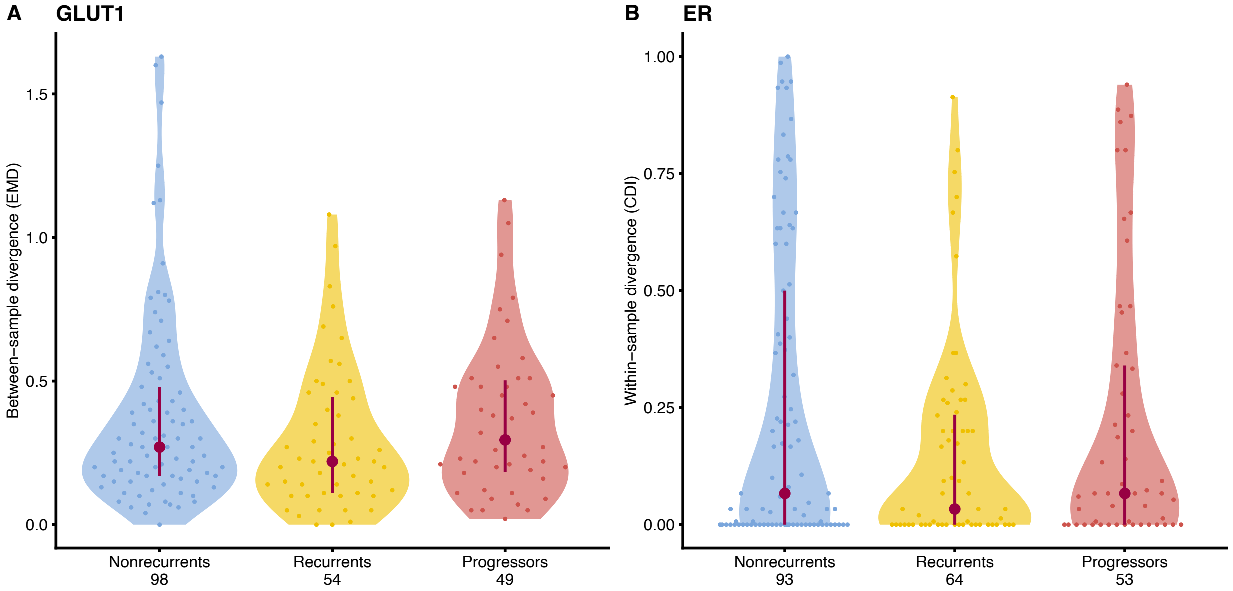


**Figure S6. Longitudinal phenotypic divergence.**

Distribution of measures of divergence per patient and marker. A, GLUT1 between-sample divergence, Earth Mover's Distance (EMD). B, ER within-sample divergence, Cumulative Density Index (CDI). Omnibus test: Kruskal-Wallis Rank Sum, Post-hoc test: Dunn's test with control for multiple tests using the Holm-Šidák adjustment. Statistically significant differences between groups shown if adjusted $p \leq 0.05$. Interquartile range (vertical line) and median (point) in burgundy. N: number of patients.

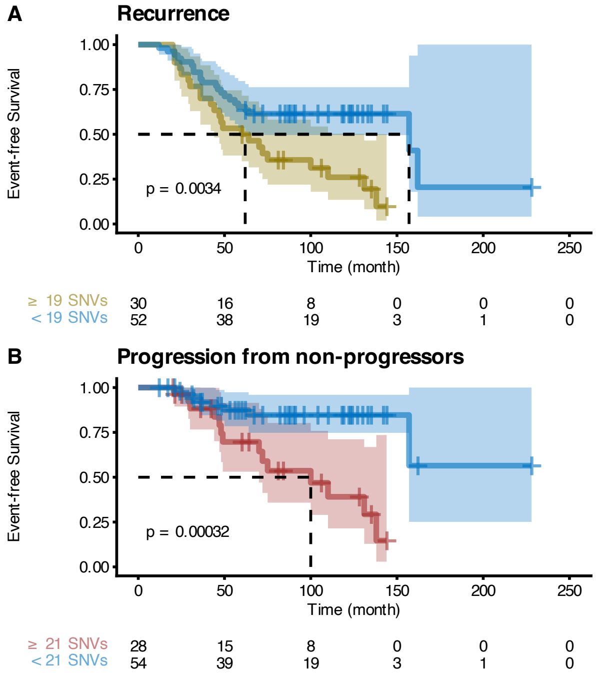


**Figure S7. Event-free survival curves of patients stratified by SNV burden: alternative clinical outcomes.**

Alternative to Fig. 6 using different clinical outcomes: recurrence instead of non-invasive recurrence and progression (right-censoring *recurrents* at time of recurrence) instead of progression (removing *recurrents*). Kaplan-Meier plots of stratified patients. **A:** Recurrence-free survival. **B:** Progression-free survival (right-censoring *recurrents* at recurrence time). SNV burden thresholds maximize Youden's J statistic of the outcomes. Log-rank test. The table below the Kaplan-Meier plot shows the number of samples at risk at different times.

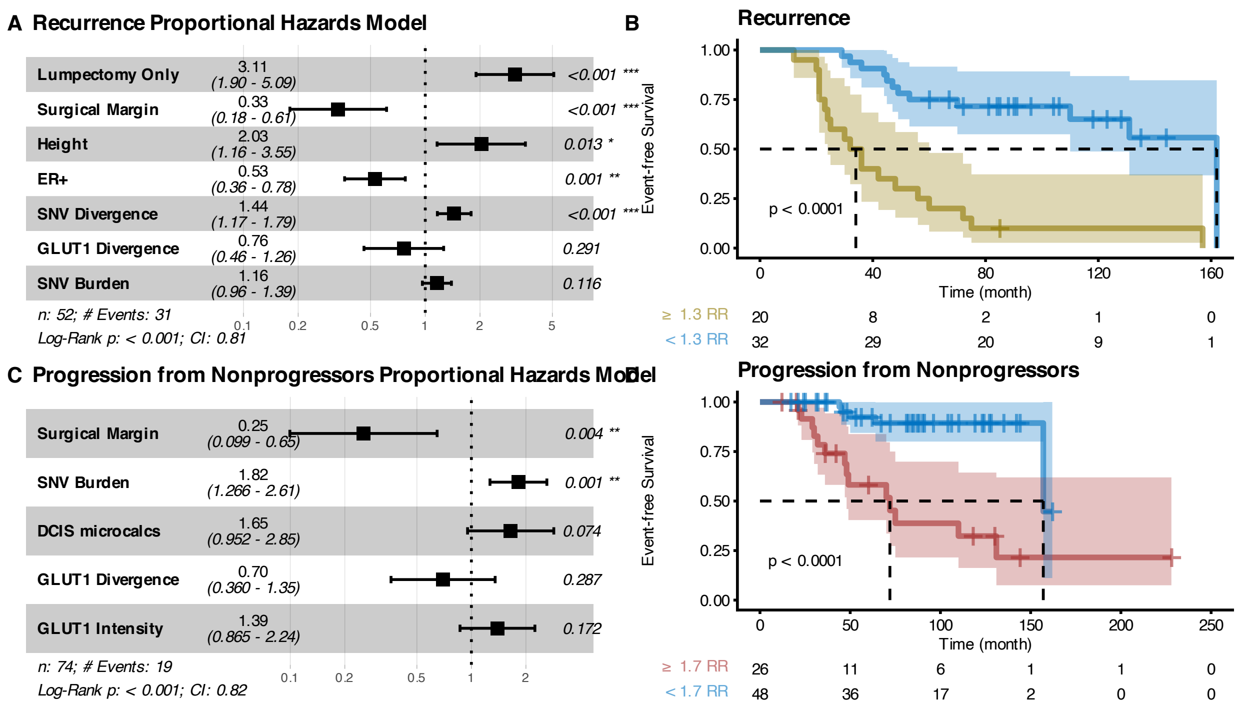


**Figure S8. Associations with time to clinical outcome: alternative clinical outcomes.** Alternative to Fig. 7 using different clinical outcomes: recurrence instead of non-invasive recurrence and progression (right-censoring *recurrents* at time of recurrence) instead of progression (removing *recurrents*). Forest plots describing proportional hazard regressions using variables selected with LASSO (**A**, **C**) and corresponding Kaplan-Meier plots of patients stratified by the relative risk threshold that maximizes Youden's J statistic of the outcomes (**B**, **D**). **A-B**: Recurrence-free survival. **C-D**: Progression-free survival (right-censoring *recurrents* at recurrence time). Log-rank test. Tables below Kaplan-Meier plots show the number of samples at risk at different times.

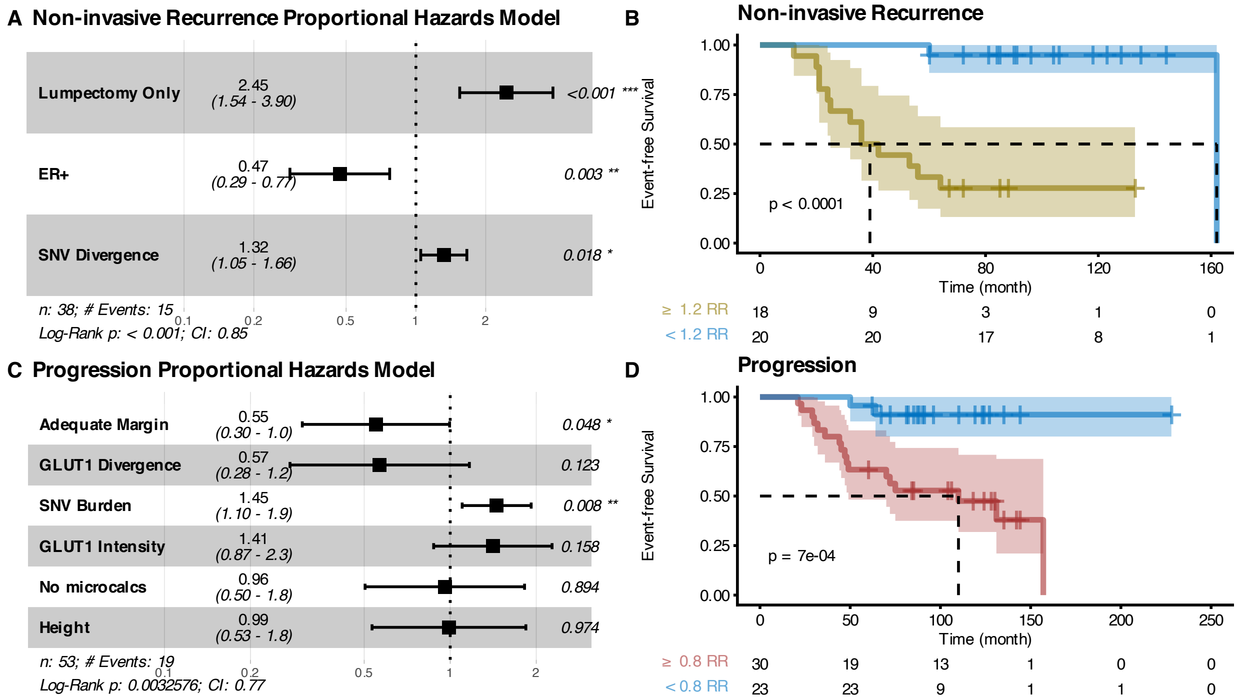


**Figure S9. Associations with time to clinical outcome: alternative clinical margin covariate.**

Alternative to Fig. 7 using a 2mm threshold to encode the surgical margin covariate. Forest plots describing proportional hazard regressions using variables selected with LASSO (**A**, **C**) and corresponding Kaplan-Meier plots of patients stratified by the relative risk threshold that maximizes Youden's J statistic of the outcomes (**B**, **D**). **A-B**: Non-invasive-recurrence-free survival. **C-D**: Progression-free survival. Log-rank test. Tables below Kaplan-Meier plots show the number of samples at risk at different times.

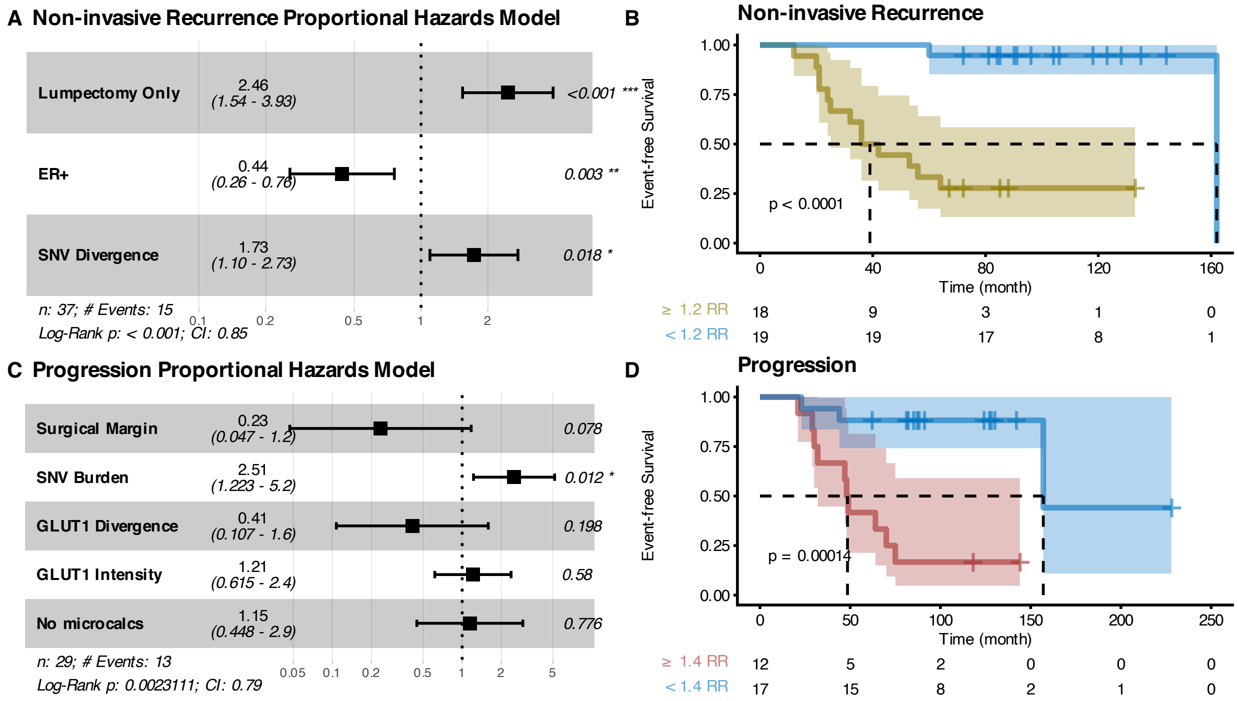


| | 0 | 40 | 80 | 120 | 160 |
|---|---|---|---|---|---|
| ≥ 1.2 RR | 18 | 9 | 3 | 1 | 0 |
| < 1.2 RR | 19 | 19 | 17 | 8 | 1 |

| | 0 | 50 | 100 | 150 | 200 | 250 |
|---|---|---|---|---|---|---|
| ≥ 1.4 RR | 12 | 5 | 2 | 0 | 0 | 0 |
| < 1.4 RR | 17 | 15 | 8 | 2 | 1 | 0 |

**Figure S10. Associations with time to clinical outcome without covariate imputation.** Forest plots describing proportional hazard regressions using variables selected with LASSO (A, C) and corresponding Kaplan-Meier plots of patients stratified by the relative risk threshold that maximizes Youden's J statistic of the outcomes (B, D). A-B: Recurrence-free survival. C-D: Progression-free survival. Log-rank test. Tables below Kaplan-Meier plots show the number of samples at risk at different times.

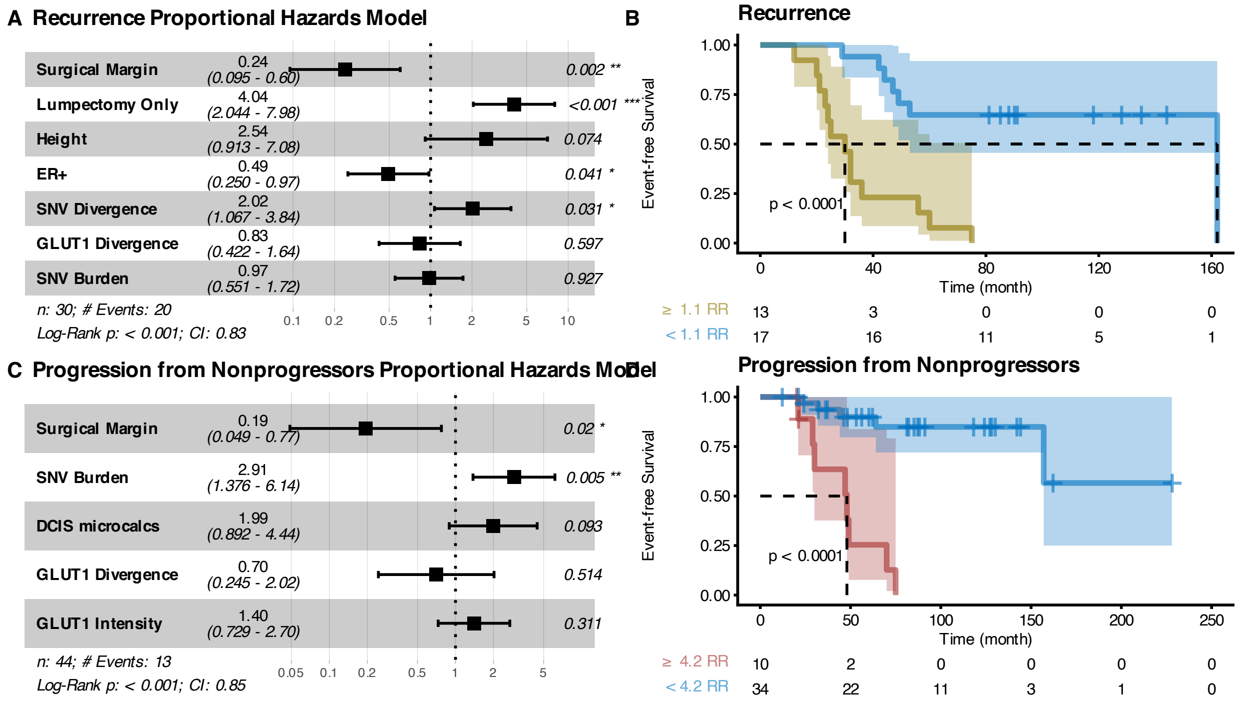


**Figure S11. Associations with time to clinical outcome without covariate imputation: alternative clinical outcomes.**

Alternative to Fig. S7 using different clinical outcomes: recurrence instead of non-invasive recurrence and progression (right-censoring *recurrents* at time of recurrence) instead of progression (removing *recurrents*). Forest plots describing proportional hazard regressions using variables selected with LASSO (A, C) and corresponding Kaplan-Meier plots of patients stratified by the relative risk threshold that maximizes Youden's J statistic of the outcomes (B, D). A-B: Recurrence-free survival. C-D: Progression-free survival (right-censoring *recurrents* at recurrence time). Log-rank test. Tables below Kaplan-Meier plots show the number of samples at risk at different times.

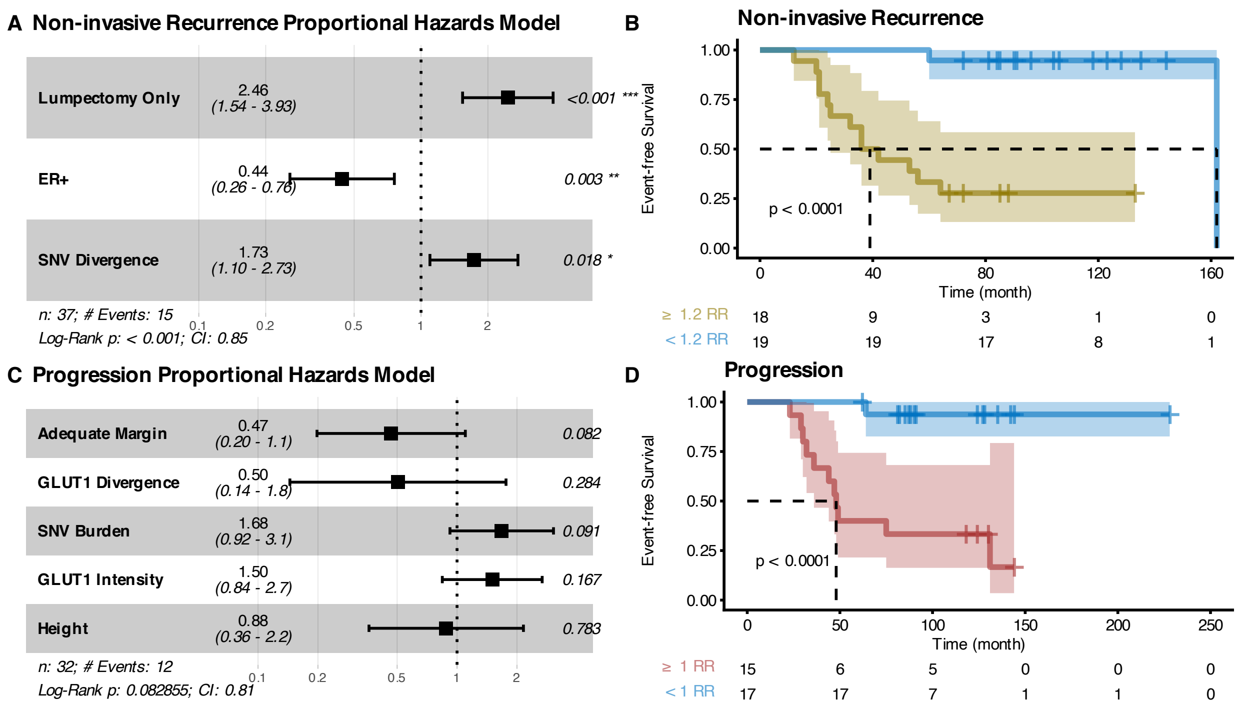


**Figure S12. Associations with time to clinical outcome without covariate imputation: alternative clinical margin covariate.**

Alternative to Fig. S7 using a 2mm threshold to encode the surgical margin covariate. Forest plots describing proportional hazard regressions using variables selected with LASSO (**A**, **C**) and corresponding Kaplan-Meier plots of patients stratified by the relative risk threshold that maximizes Youden's J statistic of the outcomes (**B**, **D**). **A-B**: Non-invasive-recurrence-free survival. **C-D**: Progression-free survival. Log-rank test. Tables below Kaplan-Meier plots show the number of samples at risk at different times.

### Supplementary Tables

**Table S1. Breakdown of the number of patients for each monoclonal immunohistochemical marker in the cross-sectional study.**

| Categories | IHC | Pure | Synchronous |
|---|---|---|---|
| Intensity (class 0-3: NULL, S, M, L) | ALDH1 | 47 | 52 |
| | CA9 | 48 | 54 |
| | ER | 42 | 48 |
| | FASN | 36 | 38 |
| | GLUT1 | 48 | 53 |
| | COX2 | 47 | 47 |
| | HER2 | 48 | 48 |
| | PFAK | 51 | 51 |
| | PR | 48 | 51 |
| | RANK | 48 | 51 |
| | COL15 | 51 | 50 |
| Signal (class 0-1: neg, pos) | ALDH1 Stroma | 47 | 51 |
| | CD68 macrophage index | 51 | 50 |
| | Ki67 | 48 | 52 |
| | FOXP3 | 44 | 52 |
| | P63 | 42 | 46 |

**Table S2. Breakdown of the number of patients for each monoclonal immunohistochemical marker in the longitudinal study.**

| Marker | Cohort | Counts |
|---|---|---|
| ER | Nonrecurrents | 92 |
| ER | Recurrents | 65 |
| ER | Progressors | 53 |
| GLUT1 | Nonrecurrents | 97 |
| GLUT1 | Recurrents | 55 |
| GLUT1 | Progressors | 49 |

**Table S3. Breakdown of the number of patients for each monoclonal immunohistochemical marker in the longitudinal study restricted to ER+ patients.**

| Marker | Cohort | Counts |
|---|---|---|
| ER | Nonrecurrents | 70 |
| ER | Recurrents | 50 |
| ER | Progressors | 43 |
| GLUT1 | Nonrecurrents | 75 |
| GLUT1 | Recurrents | 43 |
| GLUT1 | Progressors | 39 |

**Table S4. Clinical variables considered in the proportional hazard regressions of the longitudinal study.**

| Name | Type | Unit/Categories | Notes |
|---|---|---|---|
| **Age at DCIS Diagnosis** | Integer | year | |
| **Menopausal Status** | Categorical | Pre, Post | |
| **Race** | Categorical | White, Black, Other | |
| **Height** | Float | cm | |
| **Weight** | Float | kg | |
| **BMI** | Float | kg/m^2 | |
| **Axillary Dissection (at surgical treatment)** | Categorical | None, Sentinel Lymph Node Biopsy, Axillary Dissection | |
| **Number of Nodes Examined (at surgical treatment)** | Integer | number of nodes | Node fragment recoded to 1 |
| **DCIS at Surgical Margin** | Logical | | |
| **Surgical Margin** | Float | mm | Most < 10mm were measured precisely. The rest were completed from a categorical variable (<1, 1-10, >10) with 0.5, 2.5, and 10 values |
| **DCIS size** | Float | mm | |
| **Nuclear Grade** | Categorical | 1, 2, 3 | |
| **Necrosis Type** | Categorical | Comedo, non-comedo, Other, None | |
| **Microcalcification Type** | Categorical | DCIS, Benign, Both | |
| **ER** | Logical | | |
| **PR** | Logical | | |
| **DCIS Treatment** | Categorical | Lumpectomy only, Lumpectomy w Radiation, Mastectomy | |
| **Hormonal Therapy** | Logical | | |

**Table S5. DAVID functional annotation clustering results.**
Knowledgebase v2024q1. # = count, Fold = Enrichment fold.

| Nonrecurrents | | | | | | | | | | |
|---|---|---|---|---|---|---|---|---|---|---|
| **None** | | | | | | | | | | |
| **Recurrents** | | | | | | | | | | |
| **Annotation Cluster 1** | **Enrichment Score: 3.5** | | | | | | | | | |
| **Category** | **Term** | **#** | **%** | **PValue** | **Genes** | **List Total** | **Pop Hits** | **Pop Total** | **Fold** | **FDR** |
| GOTERM_MF_DIRECT | GO:0033038~bitter taste receptor activity | 4 | 5,3 | 8,2E-05 | TAS2R30, TAS2R31, TAS2R43, TAS2R46 (*Gene set 1*) | 71 | 23 | 18883 | 46,3 | 0,015 |
| INTERPRO | IPR007960:TAS2R | 4 | 5,3 | 1,3E-04 | *Gene set 1* | 74 | 28 | 20603 | 39,8 | 0,037 |
| GOTERM_BP_DIRECT | GO:0050909~sensory perception of taste | 4 | 5,3 | 1,7E-04 | *Gene set 1* | 64 | 33 | 19256 | 36,5 | 0,094 |
| “” | GO:0001580~detection of chemical stimulus involved in sensory perception of bitter taste | 4 | 5,3 | 3,0E-04 | *Gene set 1* | 64 | 40 | 19256 | 30,1 | 0,094 |
| UP_KW_BIOLOGICAL_PROCESS | KW-0919~Taste | 4 | 5,3 | 3,3E-04 | *Gene set 1* | 42 | 38 | 11447 | 28,7 | 0,007 |
| KEGG_PATHWAY | hsa04742:Taste transduction | 4 | 5,3 | 0,003 | *Gene set 1* | 31 | 86 | 8662 | 13,0 | 0,439 |
| **Progressors** | | | | | | | | | | |
| **Annotation Cluster 1** | **Enrichment Score: 3.6** | | | | | | | | | |
| **Category** | **Term** | **#** | **%** | **PValue** | **Genes** | **List Total** | **Pop Hits** | **Pop Total** | **Fold** | **FDR** |
| UP_SEQ_FEATURE | REPEAT:Spectrin 18 | 4 | 2,4 | 2,91E-05 | SPTA1, SPTBN5, DST, SPTAN1 (*Gene set 2*) | 169 | 8 | 20562 | 60,83 | 0,0110 |
| “” | REPEAT:Spectrin 19 | 4 | 2,4 | 2,91E-05 | *Gene set 2* | 169 | 8 | 20562 | 60,83 | 0,0110 |
| “” | REPEAT:Spectrin 20 | 4 | 2,4 | 2,91E-05 | *Gene set 2* | 169 | 8 | 20562 | 60,83 | 0,0110 |
| “” | REPEAT:Spectrin 15 | 4 | 2,4 | 1,12E-04 | *Gene set 2* | 169 | 12 | 20562 | 40,56 | 0,0136 |
| “” | REPEAT:Spectrin 16 | 4 | 2,4 | 1,12E-04 | *Gene set 2* | 169 | 12 | 20562 | 40,56 | 0,0136 |
| “” | REPEAT:Spectrin 17 | 4 | 2,4 | 1,12E-04 | *Gene set 2* | 169 | 12 | 20562 | 40,56 | 0,0136 |
| “” | REPEAT:Spectrin 10 | 4 | 2,4 | 1,44E-04 | *Gene set 2* | 169 | 13 | 20562 | 37,44 | 0,0136 |
| “” | REPEAT:Spectrin 11 | 4 | 2,4 | 1,44E-04 | *Gene set 2* | 169 | 13 | 20562 | 37,44 | 0,0136 |

| | | | | | | | | | | |
|---|---|---|---|---|---|---|---|---|---|---|
| “” | REPEAT:Spectrin 12 | 4 | 2,4 | 1,44E-04 | *Gene set 2* | 169 | 13 | 20562 | 37,44 | 0,0136 |
| “” | REPEAT:Spectrin 13 | 4 | 2,4 | 1,44E-04 | *Gene set 2* | 169 | 13 | 20562 | 37,44 | 0,0136 |
| “” | REPEAT:Spectrin 14 | 4 | 2,4 | 1,44E-04 | *Gene set 2* | 169 | 13 | 20562 | 37,44 | 0,0136 |
| “” | REPEAT:Spectrin 7 | 4 | 2,4 | 1,83E-04 | *Gene set 2* | 169 | 14 | 20562 | 34,76 | 0,0137 |
| “” | REPEAT:Spectrin 8 | 4 | 2,4 | 1,83E-04 | *Gene set 2* | 169 | 14 | 20562 | 34,76 | 0,0137 |
| “” | REPEAT:Spectrin 9 | 4 | 2,4 | 1,83E-04 | *Gene set 2* | 169 | 14 | 20562 | 34,76 | 0,0137 |
| “” | REPEAT:Spectrin 5 | 4 | 2,4 | 2,27E-04 | *Gene set 2* | 169 | 15 | 20562 | 32,44 | 0,0151 |
| “” | REPEAT:Spectrin 6 | 4 | 2,4 | 2,27E-04 | *Gene set 2* | 169 | 15 | 20562 | 32,44 | 0,0151 |
| “” | REPEAT:Spectrin 3 | 4 | 2,4 | 7,36E-04 | *Gene set 2* | 169 | 22 | 20562 | 22,12 | 0,0416 |
| “” | REPEAT:Spectrin 4 | 4 | 2,4 | 7,36E-04 | *Gene set 2* | 169 | 22 | 20562 | 22,12 | 0,0416 |
| INTERPRO | IPR002017:Spectrin_repeat | 4 | 2,4 | 9,18E-04 | *Gene set 2* | 167 | 24 | 20603 | 20,56 | 0,1980 |
| UP_SEQ_FEATURE | REPEAT:Spectrin 1 | 4 | 2,4 | 0,001 | *Gene set 2* | 169 | 26 | 20562 | 18,72 | 0,0596 |
| “” | REPEAT:Spectrin 2 | 4 | 2,4 | 0,001 | *Gene set 2* | 169 | 26 | 20562 | 18,72 | 0,0596 |
| INTERPRO | IPR018159:Spectrin/alpha-actinin | 4 | 2,4 | 0,002 | *Gene set 2* | 167 | 30 | 20603 | 16,45 | 0,2572 |
| SMART | SM00150:SPEC | 4 | 2,4 | 0,003 | *Gene set 2* | 110 | 29 | 10690 | 13,40 | 0,3468 |
| **Annotation Cluster 2** | **Enrichment Score: 3.3** | | | | | | | | | |
| **Category** | **Term** | **#** | **%** | **PValue** | **Genes** | **List Total** | **Pop Hits** | **Pop Total** | **Fold** | **FDR** |
| KEGG_PATHWAY | hsa05213:Endometrial cancer | 6 | 3,6 | 8,53E-05 | PIK3CA, ERBB2, PTEN, AKT1, PIK3R2, TP53 (*Gene set 3*) | 69 | 58 | 8662 | 13,0 | 0,006 |
| KEGG_PATHWAY | hsa05230:Central carbon metabolism in cancer | 6 | 3,6 | 2,10E-04 | *Gene set 3* | 69 | 70 | 8662 | 10,8 | 0,010 |
| KEGG_PATHWAY | hsa05215:Prostate cancer | 6 | 3,6 | 9,48E-04 | *Gene set 3* | 69 | 97 | 8662 | 7,8 | 0,021 |
| KEGG_PATHWAY | hsa05224:Breast cancer | 6 | 3,6 | 0,006 | *Gene set 3* | 69 | 147 | 8662 | 5,1 | 0,054 |
| **Annotation Cluster 3** | **Enrichment Score: 2.2** | | | | | | | | | |
| **Category** | **Term** | **#** | **%** | **PValue** | **Genes** | **List Total** | **Pop Hits** | **Pop Total** | **Fold** | **FDR** |
| KEGG_PATHWAY | hsa05223:Non-small cell lung cancer | 5 | 3,0 | 0,002 | PIK3CA, ERBB2, AKT1, PIK3R2, | 69 | 72 | 8662 | 8,7 | 0,036 |

| | | | | | | | | | | |
|---|---|---|---|---|---|---|---|---|---|---|
| | | | | | TP53 (*Gene set 4*) | | | | | |
| KEGG_PATHWAY | hsa05212:Pancreatic cancer | 5 | 3,0 | 0,003 | *Gene set 4* | 69 | 76 | 8662 | 8,3 | 0,038 |
| KEGG_PATHWAY | hsa05226:Gastric cancer | 5 | 3,0 | 0,029 | *Gene set 4* | 69 | 149 | 8662 | 4,2 | 0,147 |
| **Annotation Cluster 4** | **Enrichment Score: 2.1** | | | | | | | | | |
| **Category** | **Term** | | **%** | **PValue** | **Genes** | **List Total** | **Pop Hits** | **Pop Total** | **Fold** | **FDR** |
| KEGG_PATHWAY | hsa05218:Melanoma | 5 | 3,0 | 0,002 | PIK3CA, PTEN, AKT1, PIK3R2, TP53 (*Gene set 5*) | 69 | 72 | 8662 | 8,7 | 0,036 |
| KEGG_PATHWAY | hsa05214:Glioma | 5 | 3,0 | 0,003 | *Gene set 5* | 69 | 75 | 8662 | 8,4 | 0,038 |
| KEGG_PATHWAY | hsa04071:Sphingolipid signaling pathway | 5 | 3,0 | 0,015 | *Gene set 5* | 69 | 121 | 8662 | 5,2 | 0,094 |
| KEGG_PATHWAY | hsa05225:Hepatocellular carcinoma | 5 | 3,0 | 0,043 | *Gene set 5* | 69 | 168 | 8662 | 3,7 | 0,178 |

**Table S6. PANTHER overrepresentation test results using the Fisher test and FDR correction.**

| **Nonrecurrents** | | | | | | |
|---|---|---|---|---|---|---|
| PANTHER Pathways | None | | | | | |
| **Recurrents** | | | | | | |
| PANTHER Pathways | None | | | | | |
| **Progressors** | | | | | | |
| **PANTHER Pathways** | **H. sapiens REFLIST (20592)** | **Input (175)** | **Expected** | **Enrichment (fold)** | **p-val** | **FDR** |
| **Hypoxia response via HIF activation (P00030)** | 30 | 5 | .25 | 20.01 | 5.02E-06 | 8.09E-04 |
| **Hedgehog signaling pathway (P00025)** | 20 | 3 | .17 | 18.05 | 6.18E-04 | 1.66E-02 |
| **Insulin/IGF pathway-protein kinase B signaling cascade (P00033)** | 38 | 5 | .32 | 15.48 | 1.67E-05 | 8.99E-04 |
| **p53 pathway feedback loops 2 (P04398)** | 52 | 6 | .44 | 13.58 | 5.09E-06 | 4.10E-04 |
| **Axon guidance mediated by netrin (P00009)** | 35 | 3 | .30 | 10.09 | 3.23E-03 | 5.21E-02 |
| **Endothelin signaling pathway (P00019)** | 84 | 6 | .71 | 8.40 | 8.12E-05 | 3.27E-03 |
| **PI3 kinase pathway (P00048)** | 56 | 4 | .48 | 8.40 | 1.31E-03 | 2.64E-02 |
| **p53 pathway (P00059)** | 88 | 6 | .75 | 8.02 | 1.05E-04 | 3.39E-03 |
| **VEGF signaling pathway (P00056)** | 69 | 4 | .59 | 7.22 | 2.84E-03 | 5.07E-02 |
| **Ras Pathway (P04393)** | 73 | 4 | .62 | 6.45 | 3.48E-03 | 5.09E-02 |
| **EGF receptor signaling pathway (P00018)** | 138 | 6 | 1.17 | 5.12 | 1.18E-03 | 2.71E-02 |
| **Apoptosis signaling pathway (P00006)** | 119 | 5 | 1.01 | 5.34 | 3.51E-03 | 4.70E-02 |

**Table S7. Univariate proportional hazard regressions of Time to Recurrence.**

| Variable | p.val | adj.p.val | C | HR (95% CI) |
|---|---|---|---|---|
| SNV burden | 0.0022 | 0.017 | 0.58 | 1.5 (1.2 - 2) |
| CNA divergence | 0.023 | 0.16 | 0.57 | 0.75 (0.58 - 0.96) |
| ER divergence | 0.033 | 0.2 | 0.56 | 0.81 (0.66 - 0.98) |
| SNV divergence | 0.086 | 0.43 | 0.58 | 1.4 (0.96 - 1.9) |
| CNA burden | 0.12 | 0.48 | 0.54 | 1.2 (0.95 - 1.6) |
| ER intensity | 0.12 | 0.48 | 0.51 | 1.2 (0.96 - 1.4) |
| GLUT1 intensity | 0.22 | 0.48 | 0.54 | 1.1 (0.93 - 1.4) |
| GLUT1 divergence | 0.23 | 0.48 | 0.54 | 0.88 (0.71 - 1.1) |

Adj.p.val: adjusted p.value using the Holm correction. C: concordance. HR (95%CI): hazard ratio and 95% confidence interval for the standard score (z-scores) of the variable of interest.

**Table S8. Univariate proportional hazard regressions of time to progression from nonprogressors**

| Variable | p.val | adj.p.val | C | HR (95% CI) |
|---|---|---|---|---|
| SNV burden | 4.8e-05 | 0.00038 | 0.68 | 2.1 (1.5 - 3) |
| CNA burden | 0.014 | 0.096 | 0.59 | 1.5 (1.1 - 2.1) |
| GLUT1 intensity | 0.015 | 0.096 | 0.62 | 1.4 (1.1 - 1.9) |
| CNA divergence | 0.026 | 0.13 | 0.59 | 0.67 (0.48 - 0.95) |
| ER intensity | 0.076 | 0.3 | 0.54 | 1.3 (0.97 - 1.8) |
| ER divergence | 0.57 | 1 | 0.54 | 0.93 (0.71 - 1.2) |
| SNV divergence | 0.97 | 1 | 0.52 | 1 (0.59 - 1.7) |
| GLUT1 divergence | 1 | 1 | 0.51 | 1 (0.75 - 1.3) |

Adj.p.val: adjusted p.value using the Holm correction. C: concordance. HR (95%CI): hazard ratio and 95% confidence interval for the standard score (z-scores) of the variable of interest.

**Table S9. Univariate proportional hazard regressions of time to non-invasive recurrence**

| Variable | p.val | adj.p.val | C | HR (95% CI) |
|---|---|---|---|---|
| SNV divergence | 0.024 | 0.2 | 0.62 | 1.7 (1.1 - 2.8) |
| ER divergence | 0.026 | 0.2 | 0.57 | 0.72 (0.54 - 0.96) |
| CNA divergence | 0.038 | 0.23 | 0.6 | 0.68 (0.47 - 0.98) |
| GLUT1 divergence | 0.17 | 0.87 | 0.55 | 0.79 (0.57 - 1.1) |
| ER intensity | 0.19 | 0.87 | 0.52 | 1.2 (0.92 - 1.5) |
| SNV burden | 0.21 | 0.87 | 0.55 | 1.2 (0.88 - 1.8) |
| CNA burden | 0.4 | 0.87 | 0.54 | 1.2 (0.81 - 1.7) |
| GLUT1 intensity | 0.94 | 0.94 | 0.5 | 0.99 (0.77 - 1.3) |

Adj.p.val: adjusted p.value using the Holm correction. C: concordance. HR (95%CI): hazard ratio and 95% confidence interval for the standard score (z-scores) of the variable of interest.

#### Table S10. Univariate proportional hazard regressions of time to progression

| Variable | p.val | adj.p.val | C | HR (95% CI) |
|---|---|---|---|---|
| SNV burden | 7.1e-05 | 0.00057 | 0.67 | 2.2 (1.5 - 3.2) |
| CNA divergence | 0.025 | 0.17 | 0.59 | 0.68 (0.48 - 0.95) |
| ER intensity | 0.025 | 0.17 | 0.56 | 1.4 (1 - 1.9) |
| GLUT1 intensity | 0.027 | 0.17 | 0.61 | 1.4 (1 - 1.9) |
| CNA burden | 0.045 | 0.18 | 0.57 | 1.4 (1 - 2) |
| ER divergence | 0.42 | 1 | 0.54 | 0.89 (0.67 - 1.2) |
| GLUT1 divergence | 0.78 | 1 | 0.53 | 0.96 (0.7 - 1.3) |
| SNV divergence | 0.97 | 1 | 0.48 | 0.99 (0.59 - 1.7) |

Adj.p.val: adjusted p.value using the Holm correction. C: concordance. HR (95%CI): hazard ratio and 95% confidence interval for the standard score (z-scores) of the variable of interest.
